# Epidemiology of SARS-CoV-2 infection in Italy using real-world data: methodology and cohort description of the second phase of web-based EPICOVID19 study

**DOI:** 10.1101/2022.01.10.22268897

**Authors:** Fulvio Adorni, Nithiya Jesuthasan, Elena Perdixi, Aleksandra Sojic, Andrea Giacomelli, Marianna Noale, Caterina Trevisan, Michela Franchini, Stefania Pieroni, Liliana Cori, Claudio M Mastroiannii, Fabrizio Bianchi, Raffaele Antonelli Incalzi, Stefania Maggi, Massimo Galli, Federica Prinelli, on behalf of the EPICOVID19 Working Group

## Abstract

Digital technologies have been extensively employed in response to the SARS-CoV-2 pandemic worldwide. This study describes the methodology of the two-phase internet-based EPICOVID19 survey, and the characteristics of the adult volunteers respondents who lived in Italy during the first (April - May 2020) and the second wave (January - February 2021) of the epidemic. Validated scales and ad-hoc questionnaires were used to collect socio-demographic, medical and behavioural characteristics, as well as information on COVID-19. Among those who provided email addresses during phase I (105,355), 41,473 participated in phase II (mean age 50.7 years ± 13.5 SD, 60.6% females). After a median follow-up of ten months, 52.8% had undergone naso-pharyngeal swab (NPS) testing and 13.2% had positive result. More than 40% had undergone serological test (ST) and 11.9% were positive. Out of the 2,073 participants with at least one positive ST, 72.8% had only negative results from NPS or never performed it. These results indicate that a large fraction of individuals remained undiagnosed, possibly contributing to the spread of the virus in the community. Participatory online surveys offer a unique opportunity to collect relevant data at individual level from large samples during confinement.

## 1. Introduction

The coronavirus disease (COVID-19) caused by the severe acute respiratory syndrome coronavirus-2 (SARS-CoV-2), has posed an unprecedented public health emergency worldwide [1]. As of December 20, 2021, with 5,389,155 confirmed cases and 135,641 deaths Italy was the first Western country to be severely affected by the COVID-19 pandemic [2].

During the first wave of the pandemic peak worldwide, epidemiological surveillance strategies were mainly based on the testing of symptomatic patients with serious diseases requiring hospitalization and intensive medical care [3,4]. Despite efforts to ensure universal access to molecular testing, the massive spread of the infection has *de facto* restricted the diagnosis of COVID-19 to the fraction of infected people who exhibited severe symptoms only. This limitation, combined with the lack of official standards in the detection and diagnosis of asymptomatic or pauci-symptomatic patients, heavily affected the effectiveness of testing strategies and contact tracing, which in turn compromised the control of the spread of SARS-CoV-2 in the community [5]. As result of the limited availability of population-based data, the inconsistency between official statistics of different countries has made a global comparison difficult [6].

To easily and freely collect real-time and population-based data, multiple eHealth technologies have been employed [7]. In several countries, such as the UK [8], US [9], Israel [10], and Canada [11,12], large numbers of participants were recruited via mobile applications and web-based tools, to collect information on symptoms, psychosocial determinants, behavioural changes, to monitor positive cases and in some circumstances to carry out contact tracing.

The results of participatory surveillance platforms have contributed to increasing knowledge of the characteristics of SARS-CoV-2 infection and associated factors at the population level, especially in areas with insufficient testing capacity. Typical symptom patterns like anosmia, dysgeusia, fever, shortness of breath, and cough were consistently observed in association with the self-reported positive SARS-CoV-2 test, highlighting the relevance of collaborative syndromic surveillance during pandemic waves worldwide [13]. Furthermore, digital epidemiological surveillance has filled in the gaps due to the lack of seroprevalence studies, attempting to size up more completely the real, yet unknown, spread of the epidemic.

In response to the COVID-19 pandemic and to the lack of Italian epidemiological data on persons who experienced the mild-to-severe disease in the general population, a large sample of more than 198,000 voluntary adults who lived in Italy during the first lockdown was recruited through a web-based approach. These data allowed to better understand the association of symptoms (or cluster of symptoms) [14-16] and smoking habits [17] with COVID-19, the role of vaccination for other vaccine-preventable diseases [18,19], as well as to characterize psychological aspects of the population [20] and health policy issues [21] in the context of the pandemic. During the second wave of the epidemic in Italy, a follow-up questionnaire was sent by e-mail to collect further data on SARS-CoV-2 testing, COVID-19 related symptoms, hospitalization, and behavioural and psychosocial factors associated with the pandemic. This article describes the rationale, methodology, and socio-demographic and clinical characteristics of people who participated in the second phase of the internet-based EPICOVID19 study in Italy, in January-February 2021.

## 2. Methods

### 2.1 Development of the EPICOVID19 questionnaires

EPICOVID19 is an Italian national internet-based survey with a cross-sectional research design in phase I [14] and a longitudinal design in phase II, carried out on a self-selected sample of adult volunteers (18+ years old) living in Italy during the first and second waves of the pandemic. The EPICOVID19 study was established as a collaborative project of a working group including epidemiologists, physicians with expertise in infectious diseases, biostatisticians and public health professionals, with the aim of improving knowledge about SARS-CoV-2 infection. The EPICOVID19 survey was designed after a comprehensive literature review of existing research as to ensure maximal harmonisation and comparability with other large population studies. Most of the items in the questionnaire were chosen based on standardized, validated scales. The working group tested both questionnaires for two weeks and then edited them according to the feedback before launching them in the general population.

### 2.2 Content of the EPICOVID19 questionnaire

Participants were asked to complete the two questionnaires (phase I and II) after reading an introductory page (which briefly described the rationale and objectives of the study and the scientific consortium), and after accepting the option to provide consent to participate. The content of the first questionnaire was previously described in detail [14]. The phase II questionnaire is reported as Annex 1, and a comparison of its content with the one of phase I is presented in Table S1. The validated scales and questionnaires used in the two surveys are described as Table S2.

### 2.3 Sample recruitment and study population

The two-wave web-based surveys were implemented using the European Commission’s official open-source management tool EUSurvey (https://ec.europa.eu/eusurvey). The link to the first questionnaire was shared since 13^th^ April to 2^nd^ June 2020, when the Italian government was applying the strictest lockdown on the entire population. Participation was asked through mailing lists, social media platforms (Facebook, Twitter, Instagram, WhatsApp), press releases, internet pages, television and radio news programs, word of mouth and the study website (https://epicovid19.itb.cnr.it/). Inclusion criteria were age ≥18 years, access to a mobile phone, computer or tablet with internet connectivity and provision of online consent to participate in the study. In total, 207,341 participants clicked on the first questionnaire link and 198,822 provided consent to participate and completed the first online survey. Participants who had consented to be contacted (N=105,355, 53%), by providing their personal email address during the first survey, received an email invitation (since January 15 to February 28, 2021) containing a personalised link that allowed them to complete the second questionnaire. In that period the restrictions in Italy were less severe than during the first phase of the survey. Those who had not completed the EPICOVID19 phase II questionnaire within fifteen days since invitation received up to three reminder emails. Exclusion of participants who did not receive the invitation or did not respond (N=63,203), who did not provide consent (N=653), and of those with inconsistencies in email contacts or who answered more than once using the same email address (N=26) resulted in 41,473 respondents included in the present analysis (Figure 1).

**Figure 1.**
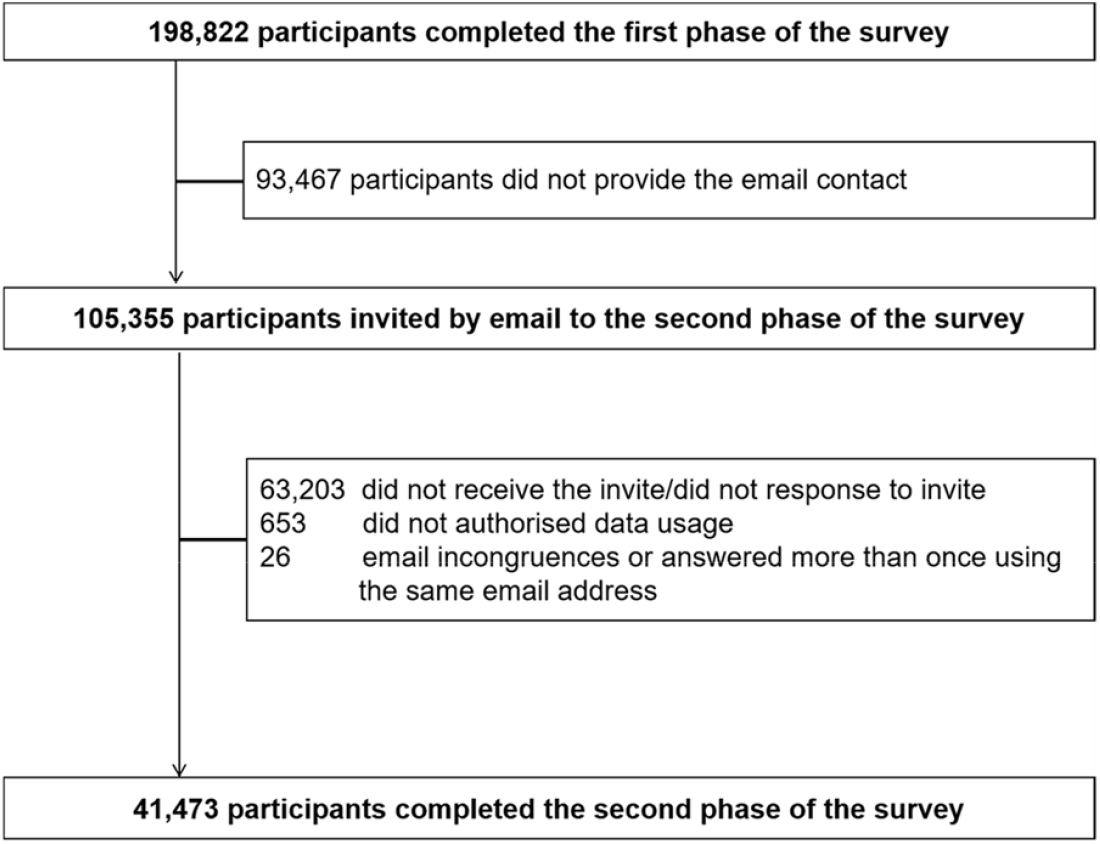
Flow-chart of the participants in phase II EPICOVID19 survey.

Excluded participants (N=157,349) were younger, more likely residents in Southern regions or islands, with a lower educational level and more frequently students (Table S3).

### 2.4 Variables collected and data transformations

Variables of interest for the present study were the following: socio-demographic information (age, education, employment, job position at-risk for the infection, socio-economic status), body mass index (BMI, calculated as weight divided by height squared), number of chronic diseases, smoking habit, alcohol consumption, self-perceived health status [22] recoded as bad or very bad, adequate, and good or very good. Townsend Deprivation Scores (TDSs) was calculated as proxy for individual level deprivation [23] by summing up, for each participant, the following variables (both dichotomized): unemployment, non-ownership of the house where he/she lives, no car owned by family members, and house crowding (defined as number of cohabitants greater than the number of rooms in the house, kitchen and bathrooms excluded). The total score ranged 0 to 4, with higher score indicating higher deprivation. Sleep problems were measured using the Jeskin Sleep Scale (JSS) [24] based on four items. Each one was rated on a Likert-like scale from 0 to 5, and the total score was the sum of all four items’ scores and ranged from 0 (no sleep problems) to 20. The continuous score was dichotomized as follows: score lower than 11 showing low frequency of sleep disturbances and score greater than 10 indicating high frequency of sleep disturbances) [25]. Personal stress was measured using the 10-item Perceived Stress Scale (PSS) [26] and adding five items developed ad-hoc. Each item was rated on a Likert-like scale 0 to 4. The score was obtained firstly by reversing responses (0 = 4, 1 = 3, 2 = 2, 3 = 1 & 4 = 0) to the four positively stated items (items 4, 5, 7, & 8) and then summing across all scale items. Individual scores fell in the range 0-40, higher scores indicating higher perceived stress. The score was categorized as follows: 0-13: low stress; 14-26: moderate stress; 27-40 high stress. Fear of contagion for oneself or relatives, fear about personal economic and job situation, and fear about the relatives’ economic and job situation were assessed with a short questionnaire developed ad-hoc for the present survey. Each aspect was rated on a Likert-like scale from 0 (no fear) to 4, and the total score was the sum of all four items’ scores and ranged from 0 to 16, with higher scores indicating higher fear. Individual feelings about being sufficiently informed about COVID-19 was dichotomized in a binary variable.

COVID-19-related variables have been reported including: contacts with COVID-19 cases, self-isolation, nasopharyngeal swab test (NPS) (numbers, results, reasons for having performed the positive test, places attended before the positive test), hospitalization, serological test (ST) (results, reasons for having performed the positive test), anti-COVID-19 vaccine, and SARS-COV-2 infection related symptoms.

### 2.5 Statistical analysis

The continuous variables were represented as mean and standard deviation (SD) and the categorical variables were expressed as numbers and percentages. The Student t-test and chi-square test were used to compare the respondents’ characteristics by sex for continuous and categorical variables, respectively. The threshold of statistical significance for any test was set at *P*-values of 0.05. All of the statistical analyses were carried out using STATA software packages (version 15, StataCorp LP, 347 College Station, Texas, USA) and SPSS (IBM Corp. Released, IBM SPSS Statistics version 25.0 Armonk, NY: IBM Corp.). The response rate map was drawn using the open-source data visualization Datawrapper GmbH tool (https://app.datawrapper.de/signin).

### 2.6 Ethical Approval

The Ethics Committee of the Istituto Nazionale per le Malattie Infettive IRCCS Lazzaro Spallanzani approved the EPICOVID19 first (protocol No. 70, 12/4/2020) and second phase (protocol No. 249, 14/1/2021) study protocols. When participants first accessed the web-based platform, they were informed about the study and its purpose, the data to be collected, and the methods of storage; they then filled in the informed consent form. Participants were able to start the EPICOVID19 questionnaire only after consenting. Participation was voluntary and no compensation was given to respondents. The planning, conduct, and reporting of the study were in line with the Declaration of Helsinki, as revised in 2013. Data were handled and stored following the European Union General Data Protection Regulation (EU-GDPR) 2016/679, and housed in the ITB-CNR server in Italy. Data transfer was safeguarded by encrypting and decrypting data and password protection. Study design and data were registered in ClinicalTrials.gov, NCT04471701.

### 2.7 Dissemination and provision of results to participants

The results of the first phase of the EPICOVID19 web-based survey were communicated mainly through peer-reviewed publications [14-21] in international scientific journals, meetings and conference presentations, workshops, the study website (www.epicovid19.itb.cnr.it); and disseminated through audio and video interviews and local printed media. A personalized e-mail with the provision of the results was sent to each participant who completed the survey and accepted to be contacted for communications about the project.

## 3. Results

The standardized response rates per 100,000 inhabitants by Italian regions over the January-February 2021 study period are represented in Figure 2 and Table S4. The percentages relating to the regional distribution of the Italian population were taken from the ISTAT website [27]. The darkest colour means the highest response rate, which was higher in the northern regions (Lombardia 137.5, Piemonte 106.6, Emilia-Romagna 100.6).

**Figure 2.**
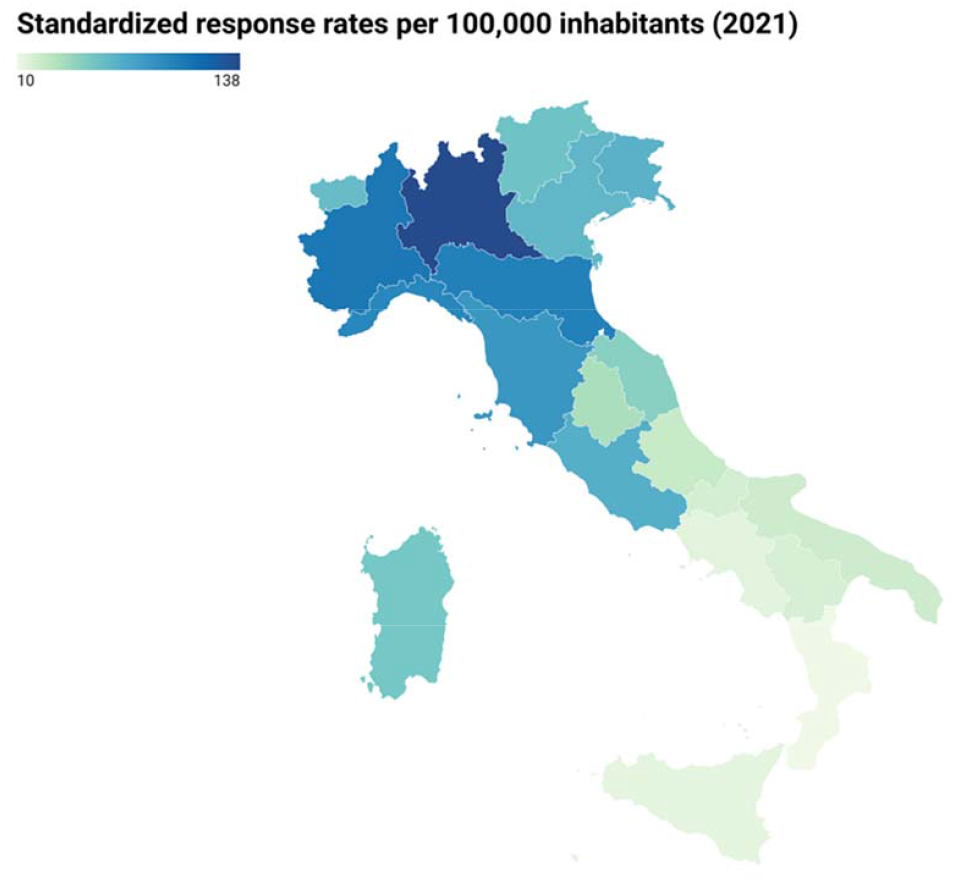
Response rates by Italian region in the phase II survey.

Table 1 summarizes the personal characteristics of the 41,473 participants who completed the phase II according to sex. The mean age of the sample was 50.7 years ± 13.5 SD (females 49.8 ± 13.0; males 52.2 ± 14.3) and 65.5% (N=27,158) had a university degree or post-graduate qualification.

**Table 1.** Individual characteristics of the study participants by sex (N=41,473).

|  | Sex at birth |  |  | Total<br>N (%) |
| --- | --- | --- | --- | --- |
|  | Females | Males | P-value |  |
|  | N (%) | N (%) |  |  |
| Age (mean ± SD) | 49.8 ± 13.0 | 52.2 ± 14.3 | .000 | 50.7 ± 13.5 |
| Class of age |  |  |  |  |
| 19-29 | 1,700 (6.8) | 1,027 (6.3) | .000 | 2,727 (6.6) |
| 30-39 | 4,434 (17.6) | 2,620 (16.0) |  | 7,054 (17.0) |
| 40-49 | 5,815 (23.1) | 3,209 (19.7) |  | 9,024 (21.8) |
| 50-59 | 6,802 (27.1) | 3,905 (23.9) |  | 10,707 (25.8) |
| 60-69 | 5,017 (20.0) | 3,628 (22.2) |  | 8,645 (20.8) |
| 70-79 | 1,262 (5.0) | 1,707 (10.5) |  | 2,969 (7.2) |
| 80+ | 116 (0.5) | 231 (1.4) |  | 347 (0.8) |
| Educational level<sup>a</sup> |  |  | .000 |  |
| Low | 723 (2.9) | 658 (4.0) |  | 1381 (3.3) |
| Middle | 7,286 (29.0) | 5,648 (34.6) |  | 12,934 (31.2) |
| High | 17,137 (68.2) | 10,021 (61.4) |  | 27,158 (65.5) |
| Employment status |  |  |  |  |
| Employed, stable position | 15,676 (62.3) | 10,448 (64.0) | .000 | 26,124 (63.0) |
| Employed, occasional worker | 1,056 (4.2) | 407 (2.5) |  | 1,463 (3.5) |
| Temporary layoff | 379 (1.5) | 114 (0.7) |  | 493 (1.2) |
| Unemployed, as before Jun 2020 | 1,243 (4.9) | 271 (1.7) |  | 1,514 (3.7) |
| Unemployed, I lost employment since Jun 2020 | 446 (1.8) | 193 (1.2) |  | 639 (1.5) |
| Student | 850 (3.4) | 583 (3.6) |  | 1433 (3.5) |
| Retired | 3,471 (13.8) | 3,420 (20.9) |  | 6,891 (16.6) |
| Other | 2,025 (8.1) | 891 (5.5) |  | 2,916 (7.0) |
| Working at |  |  | .000 |  |
| Workplace | 7,798 (46.6) | 4479 (41.3) |  | 12,277 (44.5) |
| Home and workplace | 6,188 (37.0) | 4270 (39.3) |  | 10,458 (37.9) |
| Home | 2,746 (16.4) | 2106 (19.4) |  | 4,852 (17.6) |
| Work category at risk for the infection |  |  | .000 |  |
| No | 10,242 (61.2) | 8,331 (76.7) |  | 18,573 (67.3) |
| Personnel who work indoors with high turnout | 896 (5.4) | 477 (4.4) |  | 1,373 (5.0) |
| School staff | 2,880 (17.2) | 773 (7.1) |  | 3,653 (13.2) |
| Healthcare workers | 2,572 (15.4) | 951 (8.8) |  | 3,523 (12.8) |
| Other (armed forces, haidressers, pilots, etc) | 142 (0.8) | 323 (3.0) |  | 465 (1.7) |
| Deprivation Score<sup>b</sup> |  |  | .000 |  |
| Zero | 15,005 (59.7) | 10,368 (63.5) |  | 25,373 (61.2) |
| One | 8,248 (32.8) | 4,995 (30.6) |  | 13,243 (31.9) |
| Two | 1,704 (6.8) | 893 (5.5) |  | 2,597 (6.3) |
| Three | 182 (0.7) | 68 (0.4) |  | 250 (0.6) |
| Four | 7 (0.0) | 3 (0.0) |  | 10 (0.0) |
| Body Mass Index (mean ± SD) | 23.6 ± 4.0 | 25.6 ± 3.5 | .000 | 24.4 ± 3.9 |
| N° of morbidities |  |  | .000 |  |
| None | 15,369 (61.1) | 10,660 (65.3) |  | 26,029 (62.8) |
| One | 6,042 (24.0) | 3,553 (21.8) |  | 9,595 (23.1) |
| Two | 2,416 (9.6) | 1,389 (8.5) |  | 3,805 (9.2) |
| Three or more | 1,319 (5.2) | 725 (4.4) |  | 2,044 (4.9) |
| Smoking habit |  |  | .000 |  |
| No | 14,939 (59.4) | 8,979 (55.0) |  | 23,918 (57.7) |
| Former smoker | 5,538 (22.0) | 4,545 (27.8) |  | 10,083 (24.3) |
| Current smoker | 4,669 (18.6) | 2,803 (17.2) |  | 7,472 (18.0) |
| Frequency of alcohol beverages between meals |  |  | .000 |  |
| Never | 5,739 (22.8) | 2,043 (12.5) |  | 7,782 (18.8) |
| <5 times a month | 11,409 (45.4) | 6,763 (41.4) |  | 18,172 (43.8) |
| 2-3 times a week | 4,240 (16.9) | 3,337 (20.4) |  | 7,577 (18.3) |
| 4-5 times a week | 2,033 (8.1) | 1,794 (11.0) |  | 3,827 (9.2) |
| 6+ times a week | 1,725 (6.9) | 2,390 (14.6) |  | 4,115 (9.9) |
| Self-perceived health status |  |  |  |  |
| Bad or very bad | 418 (1.7) | 221 (1.4) | .000 | 639 (1.5) |
| Adequate | 5,263 (20.9) | 2,941 (18.0) |  | 8,204 (19.8) |
| Good or very good | 19,465 (77.4) | 13,165 (80.6) |  | 32,630 (78.7) |
| Sleep problems<sup>c</sup> | 2,509 (10.0) | 850 (5.2) | .000 | 3,359 (8.1) |
| Perceived stress<sup>d</sup> |  |  | .000 |  |
| Low | 10,748 (44.1) | 9,550 (61.5) |  | 20,298 (50.9) |
| Moderate | 12,445 (51.1) | 5,633 (36.3) |  | 18,078 (45.3) |
| High | 1,168 (4.8) | 343 (2.2) |  | 1511 (3.8) |
| Fear about COVID-19 pandemic (mean ± SD)<sup>e</sup> | 8.6 ± 3.6 | 7.9 ± 3.5 | .000 | 8.4 ± 3.6 |
| Feeling to be sufficiently informed about COVID-19 | 23,443 (93.2) | 15,200 (93.1) | .607 | 38,643 (93.2) |

<sup>a</sup> Low: illiterate or primary school; middle: middle or high school; high: university or postgraduate degree
<sup>b</sup> Townsend Deprivation Score: 0 (no deprivation) to 4 (high deprivation).
<sup>c</sup> Jenkins sleep scale >12: high sleep problems.
<sup>d</sup> Perceived Stress Scale: 0-13: low stress; 14-26: moderate stress; 27-40 high stress.
<sup>e</sup> Fear score ranges from 0 (low) to 16 (high).

Respondents were mostly employed with stable positions (26,124, 63%); during the emergency period 44.5% (12,277) and 37.9% (10,458) continued to work on-site or alternated work from home and on-site work, respectively. Relatively to the risk of infection, the most represented job categories were school staff (3,653, 13.2%) and the healthcare workers (3,523, 12.8%), with significant differences between males and females. The 0.6% had a high deprivation score (score ≥3). The mean BMI was 24.4 kg/m^2^ ± 3.9 SD (females 23.6 ± 4.0; males 25.6 ± 3.5) and the 4.9% (N=2,044) of the whole sample reported three or more chronic diseases (5.2% females; 4.9% males). The 57.7% (N=23,918) were never-smokers (59.4% among females and 55.0% among males) and 25,954 (62.6%: 68.2% females and 53.9% males) were teetotallers or consumed alcoholic beverages between meals less than 5 times a month. A percentage of 8.1% reported sleep disorders during the previous month and 78.7% (N=32,630) rated his/her own health status as good or very good (77.4% females and 80.6% males). Most of the participants showed a low (50.9) or moderate (45.3) score at the PSS, with females having higher level of stress than males. More than 90% of the study participants felt they were sufficiently informed about the pandemic. Table 2 reports COVID-19-related variables according to sex. Out of all the respondents, 70.6% (N=29,300) never had close contact with COVID-19 cases and has never been in self-isolation (N=29,275). More than half of the respondents (21,877, 52.8%) underwent molecular NPS testing and among them 2,902 (13.2%) tested positive at least once, with no differences between males (13.7%) and females (13.0%).

**Table 2.**
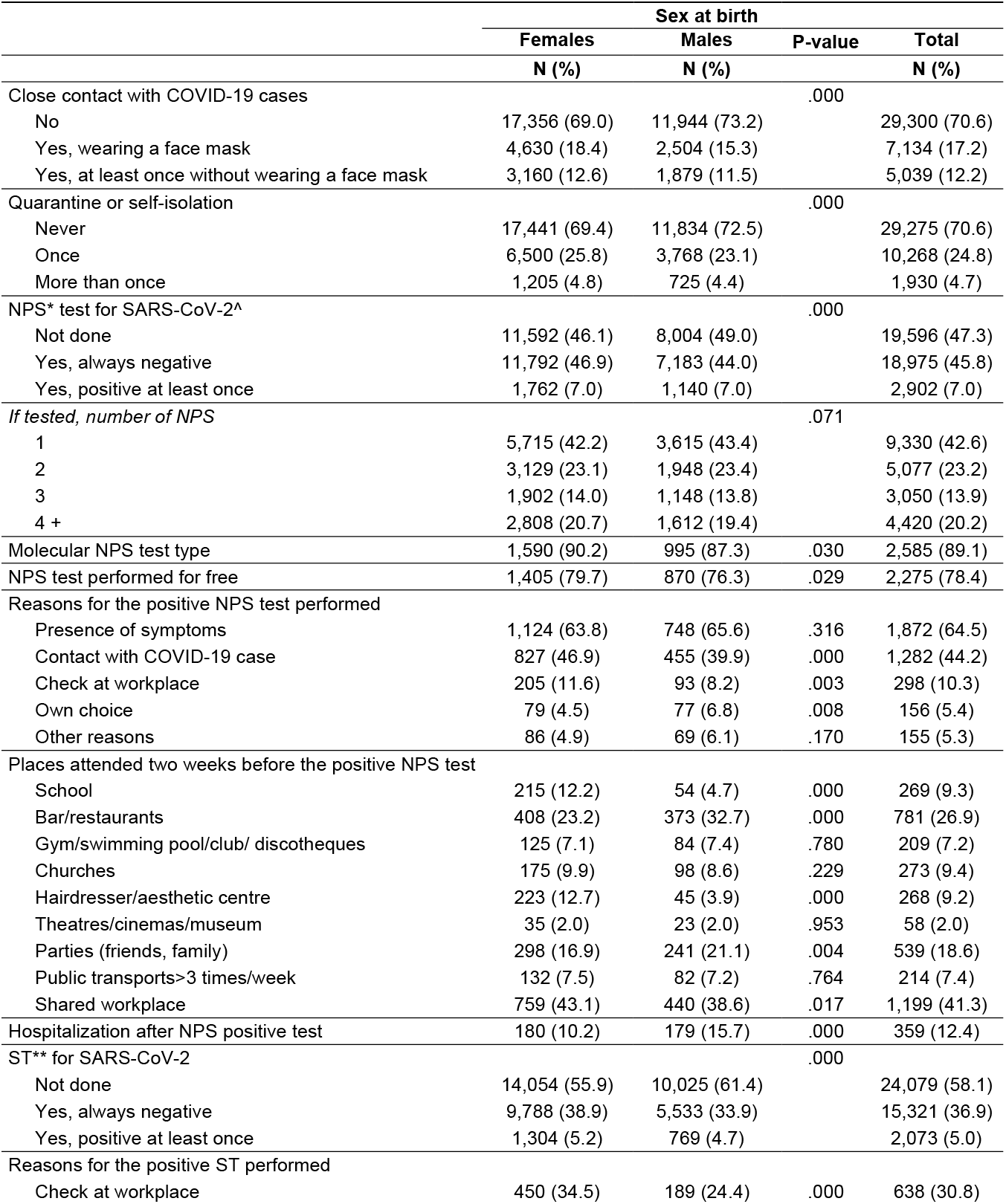
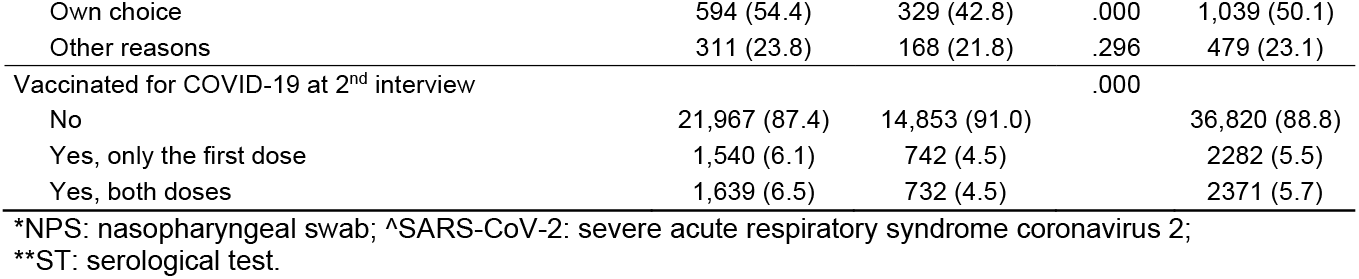
COVID-19-related variables by sex (N=41,473).

|  | Sex at birth |  | P-value | Total<br>N (%) |
| --- | --- | --- | --- | --- |
|  | Females<br>N (%) | Males<br>N (%) |  |  |
| Close contact with COVID-19 cases |  |  | .000 |  |
| No | 17,356 (69.0) | 11,944 (73.2) |  | 29,300 (70.6) |
| Yes, wearing a face mask | 4,630 (18.4) | 2,504 (15.3) |  | 7,134 (17.2) |
| Yes, at least once without wearing a face mask | 3,160 (12.6) | 1,879 (11.5) |  | 5,039 (12.2) |
| Quarantine or self-isolation |  |  | .000 |  |
| Never | 17,441 (69.4) | 11,834 (72.5) |  | 29,275 (70.6) |
| Once | 6,500 (25.8) | 3,768 (23.1) |  | 10,268 (24.8) |
| More than once | 1,205 (4.8) | 725 (4.4) |  | 1,930 (4.7) |
| NPS* test for SARS-CoV-2^ |  |  | .000 |  |
| Not done | 11,592 (46.1) | 8,004 (49.0) |  | 19,596 (47.3) |
| Yes, always negative | 11,792 (46.9) | 7,183 (44.0) |  | 18,975 (45.8) |
| Yes, positive at least once | 1,762 (7.0) | 1,140 (7.0) |  | 2,902 (7.0) |
| If tested, number of NPS |  |  | .071 |  |
| 1 | 5,715 (42.2) | 3,615 (43.4) |  | 9,330 (42.6) |
| 2 | 3,129 (23.1) | 1,948 (23.4) |  | 5,077 (23.2) |
| 3 | 1,902 (14.0) | 1,148 (13.8) |  | 3,050 (13.9) |
| 4 + | 2,808 (20.7) | 1,612 (19.4) |  | 4,420 (20.2) |
| Molecular NPS test type | 1,590 (90.2) | 995 (87.3) | .030 | 2,585 (89.1) |
| NPS test performed for free | 1,405 (79.7) | 870 (76.3) | .029 | 2,275 (78.4) |
| Reasons for the positive NPS test performed |  |  |  |  |
| Presence of symptoms | 1,124 (63.8) | 748 (65.6) | .316 | 1,872 (64.5) |
| Contact with COVID-19 case | 827 (46.9) | 455 (39.9) | .000 | 1,282 (44.2) |
| Check at workplace | 205 (11.6) | 93 (8.2) | .003 | 298 (10.3) |
| Own choice | 79 (4.5) | 77 (6.8) | .008 | 156 (5.4) |
| Other reasons | 86 (4.9) | 69 (6.1) | .170 | 155 (5.3) |
| Places attended two weeks before the positive NPS test |  |  |  |  |
| School | 215 (12.2) | 54 (4.7) | .000 | 269 (9.3) |
| Bar/restaurants | 408 (23.2) | 373 (32.7) | .000 | 781 (26.9) |
| Gym/swimming pool/club/ discotheques | 125 (7.1) | 84 (7.4) | .780 | 209 (7.2) |
| Churches | 175 (9.9) | 98 (8.6) | .229 | 273 (9.4) |
| Hairdresser/aesthetic centre | 223 (12.7) | 45 (3.9) | .000 | 268 (9.2) |
| Theatres/cinemas/museum | 35 (2.0) | 23 (2.0) | .953 | 58 (2.0) |
| Parties (friends, family) | 298 (16.9) | 241 (21.1) | .004 | 539 (18.6) |
| Public transports>3 times/week | 132 (7.5) | 82 (7.2) | .764 | 214 (7.4) |
| Shared workplace | 759 (43.1) | 440 (38.6) | .017 | 1,199 (41.3) |
| Hospitalization after NPS positive test | 180 (10.2) | 179 (15.7) | .000 | 359 (12.4) |
| ST** for SARS-CoV-2 |  |  | .000 |  |
| Not done | 14,054 (55.9) | 10,025 (61.4) |  | 24,079 (58.1) |
| Yes, always negative | 9,788 (38.9) | 5,533 (33.9) |  | 15,321 (36.9) |
| Yes, positive at least once | 1,304 (5.2) | 769 (4.7) |  | 2,073 (5.0) |
| Reasons for the positive ST performed |  |  |  |  |
| Check at workplace | 450 (34.5) | 189 (24.4) | .000 | 638 (30.8) |
| Own choice | 594 (54.4) | 329 (42.8) | .000 | 1,039 (50.1) |
| Other reasons | 311 (23.8) | 168 (21.8) | .296 | 479 (23.1) |
| Vaccinated for COVID-19 at 2<sup>nd</sup> interview |  |  | .000 |  |
| No | 21,967 (87.4) | 14,853 (91.0) |  | 36,820 (88.8) |
| Yes, only the first dose | 1,540 (6.1) | 742 (4.5) |  | 2282 (5.5) |
| Yes, both doses | 1,639 (6.5) | 732 (4.5) |  | 2371 (5.7) |

\*NPS: nasopharyngeal swab; ^SARS-CoV-2: severe acute respiratory syndrome coronavirus 2;
\*\*ST: serological test.

One fifth of tested participants performed more than four NPS during the study period and almost 90% underwent the molecular NPS test type instead of the rapid ones. The most frequent reason for the NPS testing with a positive result was the symptomatic status (64.5%) followed by having contact with a COVID-19 case (44.2%); the 41.3% referred to having shared the workplace within the 2 weeks before resulting positive to the NPS test. Among those who reported at least one positive NPS test, 359 (12.4%) were hospitalized, more frequently males (15.7%) than females (10.2%). During the study period, 41.9% of the respondents (N=17,394) underwent ST and among them 2,073 (11.9%) tested positive at least once (11.8% females and 12.2% males). Half of the participants performed the test because of their own choice (50.1%). The 5.7% of the sample (N=2,371) received both doses of the anti-COVID-19 vaccine (6.5% females and 4.5% males).

The three most frequent self-referred symptoms (Figure 3, Table S5) in the whole sample were headache (27.9%: 31.9% females and 21.6% males), sore throat/rhinorrhoea (24.5%:

**Figure 3.**
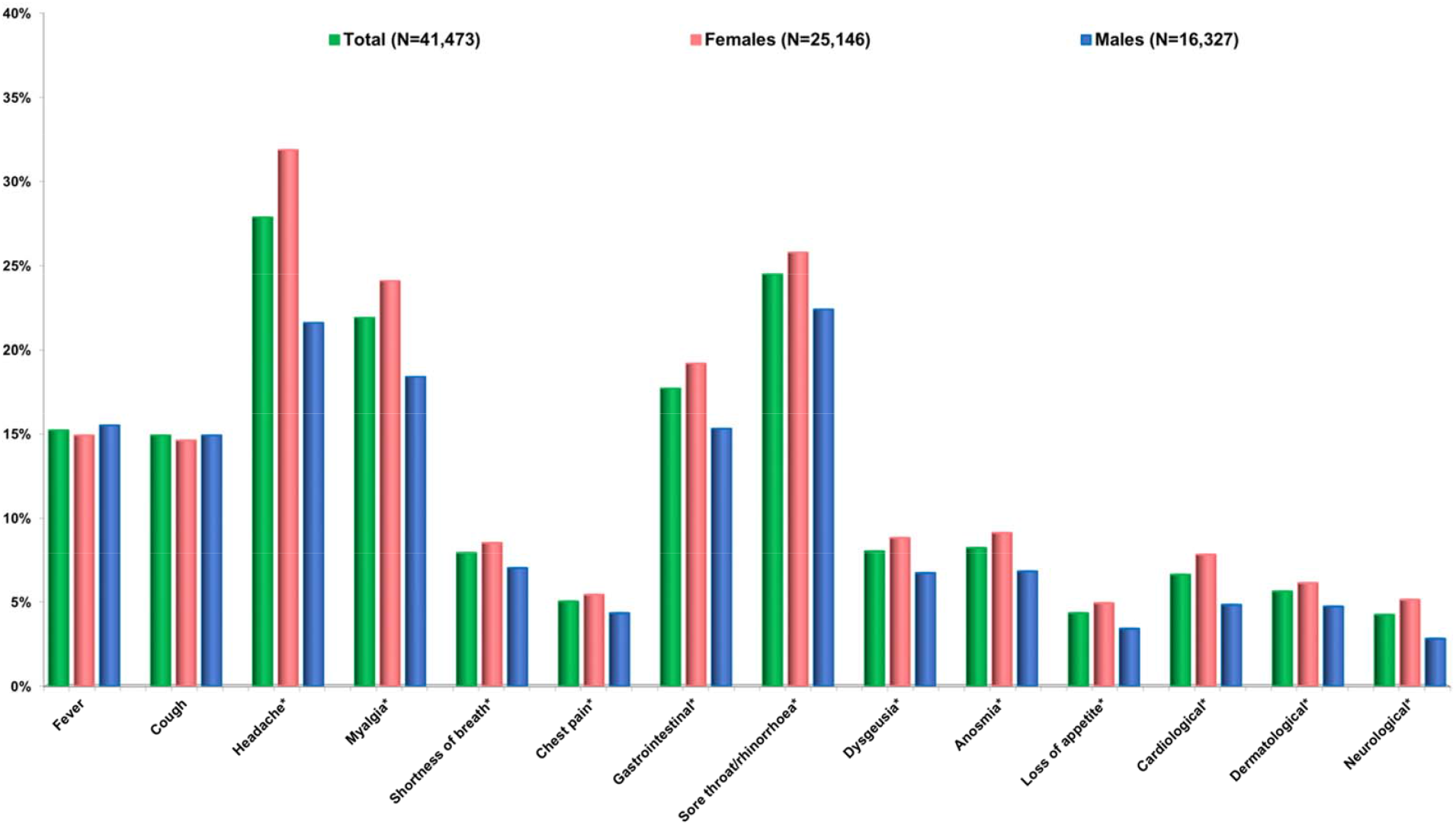
Frequency of self-reported symptoms by sex between March 2020 and February 2021 (N=41,473). *P-value < 0.05 (comparison between males and females).

25.6% females and 22.4% males), and myalgia (21.9%: 24.1% females and 18.4% males). Anosmia and dysgeusia were reported by 8.2% and 8.0% of the sample, and more frequently from females. Out of the 41,473 respondents of the second survey, 19,325 (46.6%) reported no symptoms (50.6% males and 44.0% females) (data not shown). On the other hand, among those with at least one positive NPS and/or ST (N=4,411), myalgia (64.6%: 67.6% females and 59.7% males), fever (58%: 55.4% females and 62.3% males), headache (52.7%: 58.1% females and 44% males) were the three most frequent self-reported symptoms. Anosmia and dysgeusia accounted for 51.6% (females 55.9 and males 44.7%) and 48.3% (females 52.2% and males 42.0%), respectively.

During the period March 2020 – February 2021, the 33.3% (N=13,805) did not perform any COVID-19 tests, the 24.8% (N=10,274) underwent NPS only, the 14.0% (N=5,791) underwent ST only, whereas the 28.0% (N=11,603) performed both NPS and ST (data not shown). Out of the 2,073 participants with at least one positive ST, 1,509 (72.8%) had undergone one or more NPS always with negative results or never performed it. In the group of participants diagnosed (NPS or ST) with SARS-CoV-2 infection (N=4,411), more than one-third (Figure 4) was aware that they had the infection, which was not intercepted in its acute phase (NPS never executed or executed with negative result, before or after known seropositivity), with slight differences between sexes.

**Figure 4.**
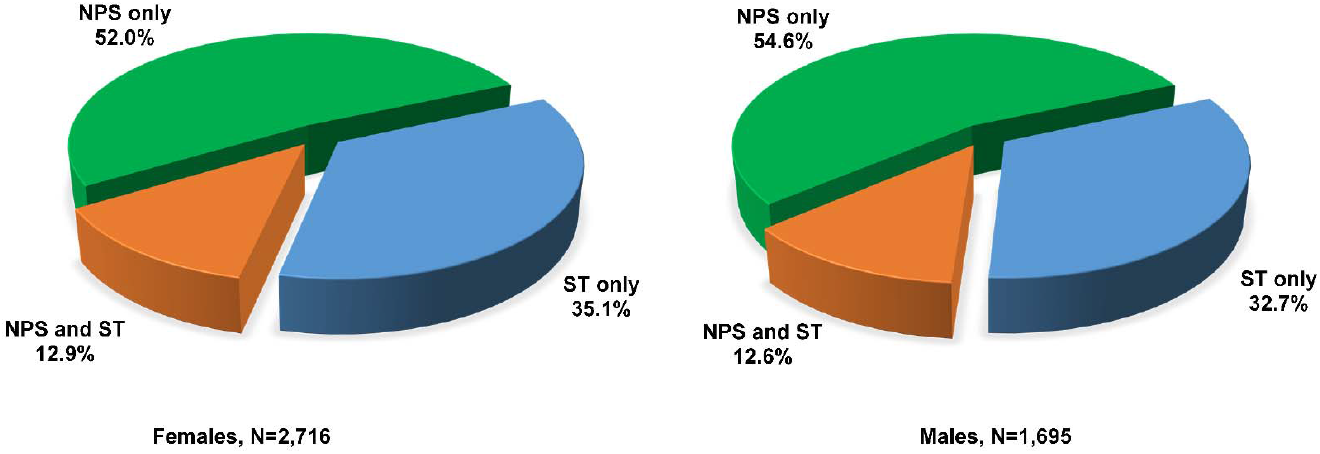
Distribution of positivity to SARS-CoV-2 by type of test performed in 4,411 cases between March 2020 and February 2021 by sex. NPS: nasopharyngeal swab. ST: serological test.

## 4. Discussion

This article provides a snapshot of the socio-demographic and clinical characteristics of the 41,473 who participated in the second phase of the web-based EPICOVID19 study conducted in Italy during January-February 2021. It also shows the frequency of the NPS and/or ST tests, the prevalence of positivity to SARS-CoV-2 among the tested participants and the frequency of COVID-19 related symptoms in a median study period of eleven months since March 2020.

The EPICOVID19 questionnaires had the power to collect some data useful to characterize the individual behaviours of the respondents involving several aspects of daily life usually not collected in clinical context. The national coverage of the survey was in line with the geographical spread of COVID-19 during the first wave [2], when participants were recruited. As to work conditions, the majority of the participants (63%) maintained their stable work position with the 18% shifting to work from home, data quite in accordance with the Eurostat Statistics. In 2020, 12.3% of employed aged 15-64 years said they often work in agile mode in the European Union, and an identical percentage was reported in Italy (12.2%) [28]. Furthermore, according to the Smart Working Observatory of the Politecnico di Milano [29], during the lockdown about 6.6 million workers shifted to remote working. Among the work category at high risk of infection, school staff and healthcare workers represent almost 30% of the study sample, with a significant unbalance toward the female sex, as expected.

Regarding the perception of health status and mood disorders, 78.7% referred to perceive a good or very good health status with no substantial difference between females and males. However, 8.1% and 50.0% reported sleep disorders and moderate-to-high self-perceived stress during the month before the survey completion, respectively. Similarly to these findings, recent studies reported a high prevalence of sleep problems [30] and a high level of stress or anxiety [31] during the COVID-19 outbreak. The results of the present study also showed that females are more likely to manifest sleep disorders and psychological stress as pointed out in other investigations [32,33]. Furthermore, females are more worried about contagion for themselves or relatives and about personal and relatives’ economic and job situation, confirming the results of phase I of the EPICOVID19 survey [20]. These data reinforce indications that although males are at higher risk of developing a severe infection than females [34], the latter are more concerned about COVID-19. This could reflect a stronger adherence to virtuous behaviours in females compared to males [35].

Considering the COVID-19-related variables, during the second survey, fever, headache myalgia, and olfactory and taste disorders were the most frequent self-reported symptoms among those who tested positive, which are consistently reported as peculiar symptoms associated with SARS-CoV-2 infection [13,14,36]. More than half of the sample underwent the NPS test (positive rate of 13.2%), because of suspected symptoms or contact with a COVID-19 case. More than 40% referred to having shared the workplace in the two weeks before having been tested. About 40% of the sample performed a ST, mostly voluntarily, and 11.9% resulted positive. Taken together, these percentages are significantly higher compared to the official number of positive cases officially reported in Italy for the period March 2020 – February 2021 (N=2.925.265 cases in 59.641.488 residents) [2,27], confirming the potential large underestimation of the actual number of exposed or infected. Ideally, only by combining large seroprevalence epidemiological studies (screening tool) with massive NPS testing (diagnostic tool) this issue could be addressed. Most recent Italian serosurveillances still report a very broad range of prevalence estimates. Vena et al. [37] reported 11% IgG and/or IgM positivity in a large adult Italian population between March and April 2020. Among the volunteers recruited in the Marche region from March to June 2020, the authors found a seroprevalence of 14.4%, without significant differences between sex and age groups [38]. As of June 2020, in a population-based study [39] carried out in a northern municipality that was heavily affected by SARS-CoV-2 infection, authors found an overall positivity to SARS-CoV-2 of 22.6%, varying according to age groups. On the other side, the Italian National Institute of Statistics (ISTAT) estimated a much lower seroprevalence of 2.5% in a large sample from 2,000 Italian municipalities during the summer of 2020 [40]. Looking at other countries, a systematic review and meta-analysis that included 47 studies involving 399,265 people from 23 countries up to 14th August 2020, reported a seroprevalence that varied from 0.37% to 22.1% in the general population. Limiting the analysis to the Italian dataset, the authors reported a pooled seroprevalence of 7.27 (95%CI 2.48–11.9) and an estimated number of people infected by SARS-CoV-2 of 4,395,587 (95%CI 1,499,457–7,249,393) [41].

Our large-scale data showed no sex difference in the proportion of respondents infected with SARS-CoV-2, in accordance with current knowledge. Although epidemiological evidence in the early phase of pandemic suggested that males had higher risk of SARS-CoV-2 infection than females [42], subsequent evidence demonstrated that this risk difference was not significant [43]. This indicates that unequal access to healthcare and testing between sexes could have skewed towards a male bias in diagnosing the infection during the first wave of the pandemic. On the other hand, males were more frequently hospitalized and possibly manifested a more severe disease than females in the present sample. This is consistent with the large body of literature reporting that males face higher rates of hospitalization, intensive therapy unit admission and death compared to females [34]. We also observed specific sex-differences in relation to the self-reported COVID-19-like symptoms, in which females tend to systematically over-report symptoms. Because no sex difference in the rate of positivity to the diagnostic or screening test has been observed, a possible explanation might reside in the fact that females were more worried about the health situation and tended to be more prone to the phenomenon of the ‘nocebo effects’ compared to males, as shown in other studies [44-46]. In line with available evidence, considering only participants with positive results from NPS and/or ST, males more often reported symptoms, such as fever and cough, known as predictors of worse outcomes [47] whereas females reported more frequently symptoms susceptible to subjective perception (headache, anosmia, dysgeusia, sore throat) and generally associated with less severe infections [48,49].

Females also completed more frequently the anti-COVID-19 vaccination cycle compared to their male counterparts. The sex unbalance can be explained by the fact that among the healthcare workers (representing 80% of those who received both vaccine doses in our sample), 73% were females (data in line with the other European Member States [50]). These results reflect the effects of the Guidelines on the Strategic Plan for COVID-19 Vaccines released on December 2^nd^, 2020 by the Ministry of Health. These Guidelines, in fact, recommended starting the vaccination campaign by first selecting specific categories, such as healthcare social workers, residents and staff working in nursing homes, at high risk of infection or of spreading the virus [51]. The low percentage of vaccinated (5.7%) in our sample was expected as the anti-COVID-19 vaccination campaign started at the beginning of January 2021 in Italy, when the survey presented in this manuscript was carried out (15 January – 28 February).

Remarkably, out of the 2,073 participants with at least one positive ST during the period March 2020–February 2021, 72.8% underwent one or more NPS always with negative results or never performed it. Among those with COVID-19, more than one-third became aware that they contracted the infection without being tested (or having a negative result at the NPS), meaning that a considerable number of undiagnosed cases escaped the detection from surveillance systems and was not officially certified as positive. COVID-19 has caught most countries unprepared and has highlighted the unreadiness of health systems [52]. In particular, during the first wave of the pandemic, most countries encountered difficulties in carrying out diagnostic tests, thus limiting the effectiveness of testing, tracking and contact tracing [5]. Consequently, the number of SARS-CoV-2 infections in the asymptomatic or subclinical infected individuals was largely undetected [53] thus leading to a considerable underestimation of the number of actual cases [54,55].

This large fraction of people, not undergone self-isolation or quarantine, is likely to have contributed to the transmission of SARS-CoV-2 infection and to the spread of the disease outbreaks in small communities such as households [56] during the most severe restrictions period, or such as workplaces [57], when the restrictions were less stringent.

The full body of results presented in this descriptive manuscript highlights a number of interesting topics that we will address in detail and with specific methodologies and approaches in future publications.

### 4.1 Limitations and Strengths

The present study has some weaknesses, primarily because the online system and voluntary participation suffers from inherent selection bias and generalizability. Similarly to other web-based survey [8,58,59], some of the characteristics of the sample were not adequately representative of the Italian adult population. Indeed, females, younger, healthier and wealthier people were proportionally more represented in the enrolled sample than in the general population. Furthermore, data were self-reported, which might have introduced measurement and recall bias (e.g. survey question misunderstanding, etc.). In addition, the longitudinal design may have led to bias due to the loss of participants during the follow-up period. The response rate to the second survey was 40%, with some differences between included and excluded participants, in particular regarding age, education, employment status and geographical area of residence.

The present study also has several strengths including its community-based longitudinal design with two-time point’s measures overlapping with the first and second wave of the epidemic in Italy, thus providing reliable details on the temporal evolution of the symptoms and testing. In addition, although these data were self-reported, almost 50% of the studied sample underwent a NPS or ST, providing an overarching picture of the positivity rate at the population level in a country in which the ability to track COVID-19 cases in real-time was limited. The exhaustive data collection on socio-demographic, medical, behavioural, and psychological factors, as well as the large sample size, is a further strength of this study.

Finally the EPICOVID19 web survey has reached a large sample of adults covering all Italian regions, although the response rate was unbalanced in favour of the northern regions, being the Italian geographical area more dramatically affected by the first wave of the pandemic at the time of enrolment.

## 5. Conclusions

EPICOVID19 is the largest web-based survey released when the first two waves of COVID-19 outbreak occurred in Italy. It offers a unique opportunity to estimate the number of individuals asymptomatic or mild symptomatic at the community level, to explore the factors associated with the SARS-CoV-2 infection, and to evaluate the consequences on health and wellbeing of the COVID-19 pandemic in Italy. The descriptive results of the phase II of the EPICOVID19 survey indicate that the positivity rate among Italian adults in February 2021 varied from 11.9% (ST) to 13.2% (NPS). Furthermore, the study highlights that a relevant fraction of positive cases remained uncertified from the official statistics, which possibly may have contributed to the spread of the virus in the community.

Complementary to the activities of testing and contact tracing, the adoption of participatory online surveys for collecting epidemiological data on a multidimensional scale should be considered strategic to support decision makers in planning evidence-based public health strategies to control the spread of SARS-CoV-2 infection.

## Supporting information

ANNEX 1 - Phase II EPICOVID19 questionnaire

## Data Availability

Data are available upon reasonable request to the authors.

## Author Contributions

Conceptualization F.A. – F.P. – M.G. – F.B. – S.M. – R.A.I.

Data Curation F.A. – F.P. – N.J. – E.P. – A.S.

Formal Analysis F.A.

Investigation F.A. – F.P.

Methodology F.A. – F.P. – A.G. – M.N. – C.T. – L.C.

Project administration F.A. – F.P.

Resources N.J. – E.P. – A.S.

Supervision F.P.

Writing – original draft F.A. – F.P.

Writing – review & editing N.J. – E.P. – A.S. – A.G. – M.N. – C.T. – M.F. – S.P. – L.C. – C.M.M. – F.B. – R.A.I. – S.M. – M.G.

## Data availability

Aggregated data and the analysis source code will be made available on reasonable request to the corresponding author.

## Acknowledgments

The authors would like to thank all the participants who took part in this study and made it possible, and all the collaborators of the EPICOVID19 Working Group (in alphabetical order by last name): Adorni Fulvio, National Research Council, Institute of Biomedical Technologies, Epidemiology Unit, Segrate (MI), Italy; Andreoni Massimo, Infectious Diseases Clinic, Department of System Medicine, Tor Vergata University of Rome, Rome, Italy; Antonelli Incalzi Raffaele, Unit of Geriatrics, Department of Medicine, Biomedical Campus of Rome, Rome, Italy; Bastiani Luca, National Research Council, Institute of Clinical Physiology, Pisa (PI), Italy; Bianchi Fabrizio, National Research Council, Institute of Clinical Physiology, Pisa (PI), Italy; Di Bari Mauro, Geriatric Intensive Care Medicine, University of Florence and Azienda Ospedaliero-Universitaria Careggi, Florence, Italy; Fortunato Loredana, National Research Council, Institute of Clinical Physiology, Pisa (PI), Italy; Galli Massimo, Infectious Diseases Unit, Department of Biomedical and Clinical Sciences L Sacco, Università di Milano, ASST Fatebenefratelli Sacco, Milan, Italy; Giacomelli Andrea, Infectious Diseases Unit, Department of Biomedical and Clinical Sciences L Sacco, Università di Milano, ASST Fatebenefratelli Sacco, Milan, Italy; Jesuthasan Nithiya, National Research Council, Institute of Biomedical Technologies, Epidemiology Unit, Segrate (MI), Italy; Maggi Stefania, National Research Council, Neuroscience Institute, Aging Branch, Padova, Italy; Mastroianni Claudio, Public Health and Infectious Disease Department, Sapienza University, Rome, Italy; Molinaro Sabrina, National Research Council, Institute of Clinical Physiology, Pisa (PI), Italy; Noale Marianna, National Research Council, Neuroscience Institute, Aging Branch, Padova, Italy; Pagani Gabriele, Infectious Diseases Unit, Department of Biomedical and Clinical Sciences L Sacco, Università di Milano, ASST Fatebenefratelli Sacco, Milan, Italy; Pedone Claudio, Unit of Geriatrics, Department of Medicine, Biomedical Campus of Rome, Rome, Italy; Pettenati Carla, National Research Council, Institute of Biomedical Technologies, Segrate (MI), Italy; Prinelli Federica, National Research Council, Institute of Biomedical Technologies, Epidemiology Unit, Segrate (MI), Italy; Rusconi Stefano, Infectious Diseases Unit, Department of Biomedical and Clinical Sciences L Sacco, Università di Milano, ASST Fatebenefratelli Sacco, Milan, Italy; Sojic Aleksandra, National Research Council, Institute of Biomedical Technologies, Epidemiology Unit, Segrate (MI), Italy; Tavio Marcello, Division of Infectious Diseases, Azienda Ospedaliero Universitaria Ospedali Riuniti, Torrette, Ancona, Italy; and Trevisan Caterina, Geriatric Unit, Department of Medicine (DIMED), University of Padova, Padova, Italy, and National Research Council, Neuroscience Institute, Aging Branch, Padova, Italy.

## References

1. European Centre for Disease Prevention and Control. Available online: https://www.ecdc.europa.eu/en/publications-data/rapid-risk-assessment-resurgence-reported-cases-covid-19 (accessed on 18 December 2021).

2. Arcgis COVID-19 Situazione Italia. Available online: http://opendatadpc.maps.arcgis.com/apps/opsdashboard/index.html#/b0c68bce2cce478eaac82fe38d4138b1 (accessed on 20 December 2021

3. Russell, T.W.; Golding, N.; Hellewell, J.; Abbott, S.; Wright, L.; Pearson, C.A.; van Zandvoort, K.; Jarvis, C.I.; Gibbs, H.; Liu, Y.; et al. Reconstructing the early global dynamics of under-ascertained COVID-19 cases and infections. BMC Med. 2020.18(1):1–9. doi: 10.1186/s12916-020-01790-9.

4. Pullano, G.; Di Domenico, L.; Sabbatini, C.E.; Valdano, E.; Turbelin, C.; Debin, M.; Guerrisi, C.; Kengne-Kuetche, C.; Souty, C.; Hanslik, T.; et al. Underdetection of cases of COVID-19 in France threatens epidemic control. Nature. 2021, 590(7844):134–139. doi: 10.1038/s41586-020-03095-6.

5. Yang, J.; Chen, X.; Deng, X.; Chen, Z.; Gong, H.; Yan, H.; et al. Disease burden and clinical severity of the first pandemic wave of COVID-19 in Wuhan, China. Nat Commun. 2020, Oct 27;11(1):5411. doi: 10.1038/s41467-020-19238-2.

6. Budd, J.; Miller, B.S.; Manning, E.M.; Lampos, V.; Zhuang, M.; Edelstein, M.; Rees, G.; et al. Digital technologies in the public-health response. Nat Med. 2020, Aug;26(8):1183–1192.

7. McCall, B. COVID-19 and artificial intelligence: protecting health-care workers and curbing the spread. Lancet Digit Health. 2020, Apr;2(4):e166–e167. doi: 10.1016/S2589-7500(20)30054-6.

8. Menni, C.; Valdes, A.M.; Freidin, M.B.; Sudre, C.H.; Nguyen, L.H.; Drew, D.A.; et al. Real-time tracking of self-reported symptoms to predict potential COVID-19. Nat Med. 2020, Jul;26(7):1037–1040. doi: 10.1038/s41591-020-0916-2.

9. Robertson, M.M.; Kulkarni, S.G.; Rane, M.; Kochhar, S.; Berry, A.; Chang, M.; Mirzayi, C.; You, W.; Maroko, A.; et al. Cohort profile: a national, community-based prospective cohort study of SARS-CoV-2 pandemic outcomes in the USA-the CHASING COVID Cohort study. BMJ Open. 2021, Sep 21;11(9):e048778.doi: 10.1136/bmjopen-2021-048778.

10. Rossman, H.; Keshet, A.; Shilo, S.; Gavrieli, A.; Bauman, T.; Cohen, O.; et al. A framework for identifying regional outbreak and spread of COVID-19 from one-minute population-wide surveys. Nat Med. 2020, May;26(5):634–638.doi: 10.1038/s41591-020-0857-9.

11. Krausz, M.; Westenberg, J.N.; Vigo, D.; Spence, R.T.; Ramsey, D. Emergency Response to COVID-19 in Canada: Platform Development and Implementation for eHealth in Crisis Management. JMIR Public Health Surveill. 2020, May 15;6(2):e18995.doi: 10.2196/18995.

12. Wu, D.C.; Jha, P.; Lam, T.; Brown, P.; Gelband, H.; Nagelkerke, N.; Birnboim, H.C.; Reid, A.; et al. Predictors of self-reported symptoms and testing for COVID-19 in Canada using a nationally representative survey. PLoS One. 2020, Oct 21;15(10):e0240778. doi: 10.1371/journal.pone.0240778. eCollection 2020.

13. Sudre, C.H.; Keshet, A.; Graham, M.S.; Joshi, A.D.; Shilo, S.; Rossman, H.; Murray, B.; Molteni, E.; Klaser, K.; et al. Anosmia, ageusia, and other COVID-19-like symptoms in association with a positive SARS-CoV-2 test, across six national digital surveillance platforms: an observational study. Lancet Digit Health. 2021, Sep;3(9):e577–e586. doi: 10.1016/S2589-7500(21)00115-1.

14. Adorni, F.; Prinelli, F.; Bianchi, F.; Giacomelli, A.; Pagani, G.; Bernacchia, D.; Rusconi, S.; et al. Self-Reported Symptoms of SARS-CoV-2 Infection in a Nonhospitalized Population in Italy: Cross-Sectional Study of the EPICOVID19 Web-Based Survey. JMIR Public Health Surveill. 2020, Sep 18;6(3):e21866.doi: 10.2196/21866.

15. Bastiani, L.; Fortunato, L.; Pieroni, S.; Bianchi, F.; Adorni, F.; Prinelli, F.; Giacomelli, A.; et al. Rapid COVID-19 Screening Based on Self-Reported Symptoms: Psychometric Assessment and Validation of the EPICOVID19 Short Diagnostic Scale. J Med Internet Res. 2021, Jan 6;23(1):e23897. doi: 10.2196/23897.

16. Trevisan, C.; Noale, M.; Prinelli, F.; Maggi, S.; Sojic, A.; Di Bari, M.; Molinaro, S.; Bastiani, L.; Giacomelli, A.; Galli, M.; Adorni, F.; et al. Age-Related Changes in Clinical Presentation of Covid-19: the EPICOVID19 Web-Based Survey. J Med Internet Res. 2021, Jan 6;23(1):e23897.

17. Prinelli, F.; Bianchi, F.; Drago, G.; Ruggieri, S.; Sojic, A.; Jesuthasan, N.; Molinaro, S.; Bastiani, L.; et al. Association Between Smoking and SARS-CoV-2 Infection: Cross-sectional Study of the EPICOVID19 Internet-Based Survey. JMIR Public Health Surveill. 2021, Apr 28;7(4):e27091. doi: 10.2196/27091.

18. Noale, M.; Trevisan, C.; Maggi, S.; Antonelli Incalzi, R.; Pedone, C.; Di Bari, M.; Adorni, F.; Jesuthasan, N.; et al. The Association between Influenza and Pneumococcal Vaccinations and SARS-Cov-2 Infection: Data from the EPICOVID19 Web-Based Survey. Vaccines. 2020, Aug 23;8(3):471. doi: 10.3390/vaccines8030471.

19. Giacomelli, A.; Galli, M.; Maggi, S.; Pagani, G.; Antonelli Incalzi, R.; et al. Missed Opportunities of Flu Vaccination in Italian Target Categories: Insights from the Online EPICOVID 19 Survey. Vaccines. 2020, Nov 9;8(4):669. doi: 10.3390/vaccines8040669.

20. Cori, L.; Curzio, O.; Adorni, F.; Prinelli, F.; Noale, M.; et al. Fear of COVID-19 for Individuals and Family Members: Indications from the National Cross-Sectional Study of the EPICOVID19 Web-Based Survey. Int J Environ Res Public Health. 2021, Mar 21;18(6):3248. doi: 10.3390/ijerph18063248.

21. Trevisan, C.; Pedone, C.; Maggi, S.; Noale, M.; Di Bari, M.; Sojic, A.; et al. Accessibility to SARS-CoV-2 swab test during the Covid-19 pandemic: Did age make the difference? Health Policy. 2021, Dec;125(12):1580–1586. doi: 10.1016/j.healthpol.2021.10.002. Epub 2021 Oct 6.

22. Eurostat. Available online: https://ec.europa.eu/eurostat/statistics-explained/index.php?title=Glossary:Self-perceived_health (accessed on 18 December 2021).

23. Townsend, P.; Phillimore, P.; Beattie A. Health and deprivation. Inequality and the North. Publisher: Croom Helm Ltd, London, UK, 1987 221 pp., ISBN 0-7099-4352-0

24. Jenkins, C.D.; Stanton, B.A.; Niemcryk, S.J.; Rose, R.M. A scale for the estimation of sleep problems in clinical research. J Clin Epidemiol. 1988, 41(4):313–21. doi: 10.1016/0895-4356(88)90138-2.

25. Monterrosa-Castro, Á.; Portela-Buelvas, K.; Salguedo-Madrid, M.; Mo-Carrascal, J.; Duran-Méndez, Leidy C. Instruments to study sleep disorders in climacteric women. Sleep Sci. 2016, Jul–Sep;9(3):169–178. doi: 10.1016/j.slsci.2016.11.001. Epub 2016 Nov 18.

26. Cohen, S.; Kamarck, T.; Mermelstein, R. A global measure of perceived stress. J Health Soc Behav. 1983, PMID: 6668417

27. I.Stat. Available online: http://dati.istat.it/ (accessed on 15 November 2021).

28. Eurostat. Available online: https://ec.europa.eu/eurostat/web/products-eurostat-news/-/edn-20210517-2 (accessed on 18 December 2021).

29. Osservatori.net digital innovation. Available online: https://www.osservatori.net/it/ricerche/comunicati-stampa/smart-working-emergenza-covid19-new-normal (accessed on 18 December 2021).

30. Alimoradi, Z.; Broström, A.; Tsang, H.W.H.; Griffiths, M.D.; Haghayegh, S.; Ohayon, M.M.; Lin, C.Y.; Pakpour, A.H. Sleep problems during COVID-19 pandemic and its’ association to psychological distress: A systematic review and meta-analysis. EClinicalMedicine. 2021, Jun;36:100916. doi: 10.1016/j.eclinm.2021.100916. Epub 2021 Jun 10.

31. Lin, L.Y.; Wang, J.; Ou-Yang, X.Y.; Miao, Q.; Chen, R.; et al. The immediate impact of the 2019 novel coronavirus (COVID-19) outbreak on subjective sleep status. Sleep Med. 2021, Jan;77:348–354. doi: 10.1016/j.sleep.2020.05.018. Epub 2020 Jun 1.

32. García-Fernández, L.; Romero-Ferreiro, V.; Padilla, S.; David López-Roldán, P.; Monzó-García, M.; Rodriguez-Jimenez, R. Gender differences in emotional response to the COVID-19 outbreak in Spain. Brain Behav. 2021, Jan;11(1):e01934. doi: 10.1002/brb3.1934. Epub 2020 Dec 11.

33. Guadagni, V.; Umilta’, A.; Iaria, G. Sleep Quality, Empathy, and Mood During the Isolation Period of the COVID-19 Pandemic in the Canadian Population: Females and Women Suffered the Most. Front Glob Womens Health. 2020, Oct 23;1:585938. doi: 10.3389/fgwh.2020.585938. eCollection 2020.

34. Peckham, H.; de Gruijter, N.M.; Raine, C.; Radziszewska, A.; Ciurtin, C.; et al. Male sex identified by global COVID-19 meta-analysis as a risk factor for death and ITU admission. Nat Commun. 2020, Dec 9;11(1):6317. doi: 10.1038/s41467-020-19741-6.

35. Galasso, V.; Pons, V.; Profeta, P.; Becher, M.; Brouard, S.; Foucault, M. Gender differences in COVID-19 attitudes and behavior: Panel evidence from eight countries. Proc Natl Acad Sci U S A. 2020, Nov 3;117(44):27285–27291. doi: 10.1073/pnas.2012520117. Epub 2020 Oct 15.

36. Giacomelli, A.; Pezzati, L.; Conti, F.; Bernacchia, D.; Siano, M.; Oreni, L.; et al. Self-reported Olfactory and Taste Disorders in Patients With Severe Acute Respiratory Coronavirus 2 Infection: A Cross-sectional Study. Clin Infect Dis. 2020, Jul 28;71(15):889–890. doi: 10.1093/cid/ciaa330.

37. Vena, A.; Berruti, M.; Adessi, A.; Blumetti, P.; Brignole, M.; Colognato, R.; et al. Prevalence of Antibodies to SARS-CoV-2 in Italian Adults and Associated Risk Factors. J Clin Med. 2020, Aug 27;9(9):2780. doi: 10.3390/jcm9092780.

38. De Santi, M.; Diotallevi, A.; Brandi, G. Seroprevalence of Severe Acute Respiratory Syndrome Coronavirus-2 (SARS-CoV-2) infection in an Italian cohort in Marche Region, Italy. Acta Biomed. 2021, Jan 25;92(1):e2021070. doi: 10.23750/abm.v92i1.10847.

39. Pagani, G.; Conti, F.; Giacomelli, A.; Bernacchia, D.; Rondanin, R.; Prina, A.; et al. Seroprevalence of SARS-CoV-2 significantly varies with age: Preliminary results from a mass population screening. J Infect. 2020, Dec;81(6):e10–e12. doi: 10.1016/j.jinf.2020.09.021. Epub 2020 Sep 19

40. Ministero della Salute. Available online: http://www.salute.gov.it/imgs/C_17_notizie_4998_0_file.pdf (accessed on 18 December 2021).

41. Rostami, A.; Sepidarkish, M.; Leeflang, M.M.G.; Riahi, S.M.; Nourollahpour Shiadeh, M.; et al. SARS-CoV-2 seroprevalence worldwide: a systematic review and meta-analysis. Clin Microbiol Infect. 2021, Mar;27(3):331–340. doi: 10.1016/j.cmi.2020.10.020. Epub 2020 Oct 24.

42. Abate, B.B.; Kassie, A.M.; Kassaw, M.W.; Aragie, T.G.; Masresha, S.A. Sex difference in coronavirus disease (COVID-19): a systematic review and meta-analysis. BMJ Open. 2020, Oct 6;10(10):e040129. doi: 10.1136/bmjopen-2020-040129.

43. The Sex, Gender and COVID-19 Project, Men, sex gender and Covid-19. Available online: https://globalhealth5050.org/the-sex-gender-and-covid-19-project/ (accessed on 18 December 2021).

44. Daniali, H.; Flaten, M.A. What Psychological Factors Make Individuals Believe They Are Infected by Coronavirus 2019? Front Psychol. 2021, Apr 22;12:667722. doi: 10.3389/fpsyg.2021.667722. eCollection 2021.

45. Daniali, H.; Flaten, M.A. Experiencing COVID-19 symptoms without the disease: The role of nocebo in reporting of symptoms. Scand J Public Health. 2021, May 27:14034948211018385. doi: 10.1177/14034948211018385.

46. Aslaksen, P.M.; Myrbakk, I.N.; Høifødt, R.S.; Flaten, M.A. The effect of experimenter gender on autonomic and subjective responses to pain stimuli. Pain. 2007, Jun;129(3):260–268. doi: 10.1016/j.pain.2006.10.011. Epub 2006 Nov 28.

47. Biolè, C.; Bianco, M.; Núñez-Gil, I.J.; Cerrato, E.; et al. Gender Differences in the Presentation and Outcomes of Hospitalized Patients With COVID-19. J Hosp Med. 2021, Jun;16(6):349–352. doi: 10.12788/jhm.3594.

48. Lechien, J.R.; Chiesa-Estomba, C.M.; Place, S.; Van Laethem, Y.; et al. Clinical and epidemiological characteristics of 1420 European patients with mild-to-moderate coronavirus disease 2019. J Intern Med. 2020, Sep;288(3):335–344. doi: 10.1111/joim.13089. Epub 2020 Jun 17.

49. Ancochea, J.; Izquierdo, J.L.; Soriano, J.B. Evidence of Gender Differences in the Diagnosis and Management of Coronavirus Disease 2019 Patients: An Analysis of Electronic Health Records Using Natural Language Processing and Machine Learning. J Womens Health (Larchmt). 2021, Mar;30(3):393–404. doi: 10.1089/jwh.2020.8721. Epub 2020 Dec 16.

50. Eurostat. Available online: https://ec.europa.eu/eurostat/web/products-eurostat-news/-/DDN-20200409-2 (accessed on 18 December 2021).

51. Ministero della Salute. Available online: https://www.salute.gov.it/portale/news/p3_2_1_1_1.jsp?lingua=italiano&menu=notizie&p=dalministero&id=5208 (accessed on 18 December 2021).

52. World Health Organization. Available online: https://www.who.int/publications/i/item/WHO-UHL-PHC-SP-2021.02 (accessed on 18 December 2021).

53. Oran, D.P.; Topol, E.J. The Proportion of SARS-CoV-2 Infections That Are Asymptomatic: A Systematic Review. Ann Intern Med. 2021, May;174(5):655–662. doi: 10.7326/M20-6976. Epub 2021 Jan 22.

54. Stringhini, S.; Wisniak, A.; Piumatti, G.; Azman, A.S.; Lauer, S.A.; Baysson, H.; et al. Seroprevalence of anti-SARS-CoV-2 IgG antibodies in Geneva, Switzerland (SEROCoV-POP): a population-based study. Lancet. 2020, Aug 1;396(10247):313–319. doi: 10.1016/S0140-6736(20)31304-0. Epub 2020 Jun 11.

55. Pollán, M.; Pérez-Gómez, B.; Pastor-Barriuso, R.; Oteo, J.; Hernán, M.A.; Pérez-Olmeda, M.; et al. Prevalence of SARS-CoV-2 in Spain (ENE-COVID): a nationwide, population-based seroepidemiological study. Lancet. 2020, Aug 22;396(10250):535–544. doi: 10.1016/S0140-6736(20)31483-5. Epub 2020 Jul 6.

56. Madewell, Z.J.; Yang, Y.; Longini, I.M.; Halloran, M.E.; Dean, N.E. Household Transmission of SARS-CoV-2: A Systematic Review and Meta-analysis. JAMA Netw Open. 2020, Dec 1;3(12):e2031756. doi: 10.1001/jamanetworkopen.2020.31756.

57. Daniels, S.; Wei, H.; Han, Y.; Catt, H.; Denning, D.W.; Hall, I.; et al. Risk factors associated with respiratory infectious disease-related presenteeism: a rapid review. BMC Public Health. 2021, Oct 28;21(1):1955. doi: 10.1186/s12889-021-12008-9.

58. Drew, D.A.; Nguyen, L.H.; Steves, C.J.; Menni, C.; Freydin, M.; Varsavsky, T.; et al. Rapid implementation of mobile technology for real-time epidemiology of COVID-19. Science. 2020, Jun 19;368(6497):1362–1367. doi: 10.1126/science.abc0473. Epub 2020 May 5.

59. Astley, C.M.; Tuli, G.; >Mc Cord, K.A.; Cohn, E.L.; Rader, B.; Varrelman, T.J.; et al. Global monitoring of the impact of the COVID-19 pandemic through online surveys sampled from the Facebook user base. Proc Natl Acad Sci U S A. 2021, Dec 21;118(51):e2111455118. doi: 10.1073/pnas.2111455118.

