## Supplementary material for "Epidemiology of SARS-CoV-2 infection in Italy using real-world data: methodology and cohort description of the second phase of web-based EPICOVID19 study": ANNEX 1 - Phase II EPICOVID19 questionnaire

### QUESTIONNAIRE EPICOVID19 – PHASE 2

#### SOCIO-DEMOGRAPHIC DATA

**Mandatory questions*

1. **Sex at birth*** M |__| F |__| Prefers not to answer |__|

2. **Province of domicile*** |__|__|__|__|__|__|__|__|__|__|__|__|__|__|__|__|__|__|__|__|__|__|__|

3. **Municipality*** |__|__|__|__|__|__|__|__|__|__|__|__|__|__|__|__|__|__|__|__|__|__|__|__|__|

4. **Postal code*** |__|__|__|__|__|

5. **Indicate your employment status from the beginning of June 2020 to the beginning of the restrictive measures in November:***

|__| Employed, steady employment
|__| Employed, occasional worker (e.g. on-call work)
|__| Student
|__| Unemployed now as well as before the reference period
|__| Unemployed now because I lost my job in the meantime
|__| Retired
|__| Temporary layoff
|__| None of the above

6. *If the answer is yes to one of the first two answers:**

|__| I resumed working exclusively on-site, at my workplace
|__| I work from home only
|__| I alternate work from home with on-site work

7. **In the last 6 months (from the beginning of June 2020 to date), did your occupation include, even partially, one of the following? (multiple choices are possible)***

|__| Healthcare workers in close contact with the patient (e.g. clinician, nurses, dentists, social health operator, rescuers)
|__| School staff who works in the presence of children or teenagers (teachers, educators, cleaning staff)
|__| Personnel who work in highly crowded rooms or closed environments (e.g. supermarkets, bars, restaurants, gyms, swimming pools, shops, clubs, discos)
|__| Armed forces occupations (e.g. policeman, soldier)
|__| Bus and tram drivers, flight attendants, taxi drivers
|__| Staff working in beauty salons and hairdressers
|__| None of the above

#### CLINICAL FEATURES AND COVID-19 RELATED VARIABLES

8. **Weight*** |__|__|__| Kg

9. **Height*** |__|__|__| cm

10. **Please indicate your blood type***

|__| A
|__| B
|__| AB
|__| O
|__| I don't know

**11. Do you have or have you had one or more of the following symptoms? If yes, indicate when (more choices are possible)***

| **Symptoms** | **Yes** | **Before September 2020** | **September 2020** | **October 2020** | **November 2020** | **December 2020** | **January 2021** |
|---|---|---|---|---|---|---|---|
| Fever with a temperature greater than 37.5 °C for at least one day | | | | | | | |
| Cough | | | | | | | |
| Sore throat/rhinorrhoea | | | | | | | |
| Headache | | | | | | | |
| Myalgia | | | | | | | |
| Taste disorder | | | | | | | |
| Olfactory disorder | | | | | | | |
| Difficulty breathing, shortness of breath due to light exertion (e.g. walking or climbing a few steps) and / or shortness of breath at rest | | | | | | | |
| Chest pain (sternum pain) | | | | | | | |
| Gastrointestinal disorders (diarrhoea, nausea, vomiting) | | | | | | | |
| Dermatological manifestations (rashes) | | | | | | | |
| Loss of appetite | | | | | | | |
| Heart arrhythmia, palpitations, tachycardia, chest tightness | | | | | | | |
| Other neurological disorders (dizziness, confusion, memory problems) | | | | | | | |
| No symptoms | | | | | | | |

**12. If you have had symptoms attributable to a suspected or ascertained infection (swab and/or serological test), please indicate for each one whether:**

| **Symptoms** | **I have fully recovered** | **I partially recovered** | **I have not recovered at all** |
|---|---|---|---|
| Fever with a temperature greater than 37.5 °C for at least one day | | | |
| Cough | | | |
| Sore throat/rhinorrhoea | | | |
| Headache | | | |
| Myalgia | | | |
| Taste disorder | | | |
| Olfactory disorder | | | |
| Difficulty breathing, shortness of breath due to light exertion (e.g. walking or climbing a few steps) and / or shortness of breath at rest | | | |
| Chest pain (sternum pain) | | | |
| Gastrointestinal disorders (diarrhoea, nausea, vomiting) | | | |
| Dermatological manifestations (rashes) | | | |
| Loss of appetite | | | |
| Heart arrhythmia, palpitations, tachycardia, chest tightness | | | |
| Other neurological disorders (dizziness, confusion, memory problems) | | | |

**13. Please indicate if you have or have been diagnosed with one or more of these diseases or disorders***

| **Diseases or ailments** | **Before June 2020** | **From June 2020 to date** |
|---|---|---|
| Lung diseases (e.g. asthma, obstructive pulmonary disease, metapneumonic fibrosis) | | |
| Heart diseases (e.g. ischemic heart disease, atrial fibrillation, heart failure) | | |
| Hypertension | | |
| Kidney diseases or dialysis | | |
| Immune system diseases (e.g. celiac disease, Graves' disease, thyroid disease, psoriasis, rheumatoid arthritis, systemic lupus erythematosus, type I diabetes) | | |
| Tumours | | |
| Metabolic diseases (e.g. type II diabetes, obesity, gout) | | |
| Neurological diseases( e.g. multiple sclerosis, epilepsy, neuropathies) | | |
| Cerebrovascular diseases (e.g. stroke, ischemia) | | |
| Hepatitis B | | |
| Hepatitis C | | |

| |
|---|
| Other liver diseases (e.g. liver cirrhosis, liver failure) |
| Depression and/or anxiety |
| Eating disorders (bulimia, anorexia, or other eating disorders) |
| Anaemia |
| No disease or disorder |

**14. Please indicate if these conditions have occurred during your life***

| **Other conditions** |
|---|
| Not self-sufficient in carrying out daily activities (e.g. washing, dressing, shopping, cooking) |
| Allergies to pollens |
| Food allergy |
| Recurrent herpes |
| Cytomegalovirus infection |
| Papilloma virus infection |
| None of these conditions |

**15. Indicate if you regularly use these medications or supplements (not as a treatment for COVID-19):***

|__| Oral or injection cortisones
|__| Inhaled cortisones and/or bronchodilators
|__| Antidepressants/anxiolytics/sedatives
|__| Vitamin D (e.g. DIBASE)
|__| Multivitamins

**16. Did you have the following vaccinations?***

| **Vaccinations** |
|---|
| Anti-flu in the winter season 2020/2021 |
| Anti-pneumococcal in the past 5 years |
| Anti-zoster |
| Anti-meningococcal |
| Anti-tetanus in the past 10 years |
| None of these vaccinations |

**17. Did you get the COVID19 vaccination?***

|__| No
|__| Yes, only the first dose
|__| Yes, both doses

*To be completed only if answer 17 is "Yes, only the first dose" or "Yes, both doses"*

**18. Indicate the date of the first vaccine dose?***

|__|__| /|__||__|/|__||__||__||__|

*To be completed only if answer 17 is “No”*

**19. Thinking about the COVID-19 vaccine:**

|__| I will definitely get the vaccine
|__| I will probably get the vaccine, but before that I will inform myself better

|__| I will probably not get the vaccine, but in any case I will inform myself better
|__| I will not get the vaccine
|__| I can't answer at this moment

*****************************************************************************************************************************

*To be completed only if of female sex*

**20. Are you pregnant?***
|__| No |__| Yes |__| I'm in menopause

**21. Indicate the number of completed pregnancies***
|__| 0 |__| 1 |__| 2 or more

**22. Have you started taking contraceptives/birth pills from 1 June 2020 to date?***
|__| No
|__| Yes

**23. Have you started taking hormone replacement therapy from 1 June 2020 to date?***
|__| No
|__| Yes

*To be completed only if of male sex*

**24. Are you following or have you been following hormone therapy (e.g. anti-androgen)?***
|__| Never
|__| Yes, in the past, for less than 5 years
|__| Yes, in the past, for more than 5 years
|__| Yes currently, taking it less than 5 years
|__| Yes currently, taking it more than 5 years

*****************************************************************************************************************************

**25. Have you been in a close contact (direct contact at a distance of less than 2 meters, and lasting more than 15 minutes in a closed environment such as a house, workplace, waiting room, airplane, transportation vehicles) with confirmed COVID-19 cases, live or deceased?***
|__| No, as far as I know, I have not been in a close contact
|__| Yes, at least once without the mask
|__| Yes, at least once but always with a mask

**26. Have you ever been in fiduciary isolation (or quarantine) either self-imposed or established by the health authorities?***
|__| Never
|__| Yes, once
|__| Yes, more than once

#### NASOPHARYNGEAL SWAB FOR COVID-19

**27. Since September 1, have you had at least one swab for COVID-19?***
|__| No, I did not perform any test
|__| Yes, at least once with a positive result

|__| Yes, always with a negative result

*If the answer is "Yes" to the last two answers:*

**28. How many swab tests did you take?***
|__| 1
|__| 2
|__| 3
|__| 4 or +

*To be completed only if answer 27 is "Yes, at least once with a positive result"*

**Considering the first swab with a positive result, indicate:**

**29. Month in which it was performed*** |__|__|__|__|__|

**30. Type of swab test***
|__| Molecular/PCR (outcome after at least 24 hours)
|__| Rapid (results in less than one hour)
|__| I do not know

**31. Did you pay for the swab?***
|__| No |__| Yes

**32. Reason for swabbing (multiple choices are possible)***
|__| I had developed suspicious symptoms
|__| I have had close contact with a confirmed COVID-19 case (in the home, workplace, etc.)
|__| For workplace control (e.g. healthcare workers)
|__| Personal choice
|__| Other reasons than those listed above

*To be completed only if you have had symptoms*

**33. Number of days elapsed between the onset of symptoms and the swab test***
|__| 1-2 days
|__| 3-7 days
|__| 8-14 days
|__| More than 14 days

**34. In the two weeks before the swab, did you attend one or more of these places? (multiple choices are possible)***
|__| Schools
|__| Bars, restaurants
|__| Gyms, swimming pools, clubs, discotheques
|__| Churches, oratories
|__| Beauty center, hairdresser
|__| Theatres, cinemas, museums
|__| Parties, lunches, dinners at home
|__| Public transport more than 3 times a week
|__| Shared working spaces in less than two meters away with at least one other person

**35. Did you ever go to the hospital after the infection?***
|__| Never
|__| Yes, but I was sent back and treated at home
|__| Yes, I was hospitalized and underwent pharmacological treatments with or without oxygen in the ward with a simple mask or nasal glasses but without the need for a helmet / CPAP

|__| Yes, I was hospitalized and underwent pharmacological treatment with or without oxygen with non-invasive ventilation (e.g. helmet / CPAP)
|__| Yes, I was hospitalized and underwent to pharmacological treatments and intubation

*To be completed only in case of hospitalization*

**36. How many days did the hospitalization last?*** |__||__||__| days

**37. What treatments did you undergo for the treatment of COVID-19? (multiple choices are possible)***
|__| Low molecular weight heparin
|__| Cortisones
|__| Azithromycin
|__| Other antibiotics
|__| Antipyretics or pain relievers
|__| Remdesevir
|__| Lopinavir / ritonavir (Kaletra)
|__| Clorochina
|__| Oxygen therapy
|__| Other or do not know
|__| None

**38. Did you have at least one control swab test?***
|__| Yes, and my last swab is positive ==> Please indicate the number of days between the first positive swab and the control test | ___ || ___ |
|__| Yes, and my last swab is negative ==> Please indicate the number of days between the first positive swab and the control test | ___ || ___ |

|__| No, I haven't done it ==> Has it been more than a month since the first positive swab to date?

| __ | No | __ | Yes

*To be completed only if answer 24 is "Yes, always with a negative result"*

**If you have always swabs with negative results, think about your last negative swab and indicate:**

**39. Month in which it was made*** |__|__|__|__|__|

**40. Type of swab test***
|__| Molecular /PCR (outcome after at least 24 hours)
|__| Rapid (results in less than one hours)
|__| I do not know

**41. Did you have to pay for the swab?***
|__| No |__| Yes

**42. Reason for swabbing (multiple choices are possible)***
|__| I had developed suspicious symptoms
|__| I have had close contact with a confirmed COVID-19 case (at home, workplace, etc.)
|__| For workplace control (e.g. health care workers)
|__| Personal choice
|__| Other reasons than those listed above

TO BE COMPLETED ONLY IF HAS SYMPTOMS

**43. Number of days elapsed between the onset of symptoms and the swab test***
|__| 1-2 days
|__| 3-7 days
|__| 8-14 days

|__| More than 14 days

**44. In the two weeks before the swab, did you attend one or more of these places? (multiple choices are possible)***

|__| Schools
|__| Bars, restaurants
|__| Gyms, swimming pools, clubs, discotheques
|__| Churches, oratories
|__| Beauty center, hairdresser
|__| Theatres, cinemas, museums
|__| Parties, lunches, dinners at home
|__| Public transport more than 3 times a week
|__| Shared working spaces in less than two meters away with at least one other person

**<u>SEROLOGICAL TEST FOR COVID-19</u>**

**45. Since September 1st, have you done at least one serological test (blood drop or blood sample) for COVID-19?***

|__| No, I did not perform any test
|__| Yes, at least once with a positive result
|__| Yes, always with a negative result

*To be completed only if the answer is "Yes, at least once with a positive result"*

**46. Thinking about the first (or the only) positive serological test, indicate the month in which it was performed*** |__||__||__||__||__||__|

**47. Type of test*** |__| Lancing device |__| Blood sample

**48. Reason for doing the serological test (multiple choices are possible)***

|__| For workplace control (e.g. healthcare workers)
|__| Personal choice
|__| Other reasons than those listed above

**49. After the first positive serological test, have you ever had a negative serological test?***

|__| No, I've never done any other serological tests
|__| No, my last serological test is still positive => Last month positive test | ___ || ___ || ___ || ___ |
|__| Yes => Month of first negative test | ___ || ___ || ___ || ___ |

*To be completed only if the answer is "Yes, always with a negative result"*

**50. Thinking about the last negative serological test, indicate the month in which it was performed*** |__||__||__||__||__||__|

**51. Type of test*** |__| Lancing device |__| Blood sample

**52. Reason for doing the serological test (multiple choices are possible)***

|__| For workplace control (e.g. healthcare workers)
|__| Personal choice
|__| Other reasons than those listed above

#### QUESTIONNAIRE EPICOVID19 – PHASE 2

##### PERSONAL CHARACTERISTICS AND HEALTH STATUS

**53. How would you describe your health in general?***
|__| Very bad |__| Bad |__| Adequate |__| Good |__| Very good

**54. After the COVID-19 pandemic situation:***

| | No | A little bit | Neutral | Enough | Yes |
|---|---|---|---|---|---|
| Are you afraid of being infected with COVID-19? | | | | | |
| Are you afraid that your family members might be infected? | | | | | |
| Are you worried about your economic and working conditions? | | | | | |
| Are you worried about the economic and working conditions of your family members? | | | | | |

**55. How many days in the past 30 days did you:***

| | 0 | 1 | 2 | 3 | 4 | 5 |
|---|---|---|---|---|---|---|
| Have trouble falling asleep? | | | | | | |
| Wake up several times per night but did not have trouble falling asleep again? | | | | | | |
| Wake up one or more times per night (including waking far too early) and have trouble falling asleep again? | | | | | | |
| Wake up after your usual amount of sleep feeling tired or exhausted? | | | | | | |

*0 = Not at all 1 = 1-3 days 2 = 4-7 days 3 = 8-14 days 4 = 15-21 days 5 = 22-30 days*

**56. Thinking about the last month, how often:***

| | 0 | 1 | 2 | 3 | 4 |
|---|---|---|---|---|---|
| Have you been upset because of something that happened unexpectedly? | | | | | |
| Have you felt that you were unable to control the important things in your life? | | | | | |
| Have you felt nervous and “stressed”? | | | | | |
| Have you felt confident about your ability to handle your personal problems? | | | | | |
| Have you felt that things were going your way? | | | | | |
| Have you found that you could not cope with all the things that you had to do? | | | | | |
| Have you been able to control irritations in your life? | | | | | |
| Have you felt that you were on top of things? | | | | | |
| Have you been angered because of things that were outside of your control? | | | | | |
| Have you felt difficulties were piling up so high that you could not overcome them? | | | | | |
| Have you trusted yourself? | | | | | |
| Have you trusted others? | | | | | |
| Have you felt you can protect your loved ones? | | | | | |
| Have you felt you can help improve the situation? | | | | | |
| Have you felt helpless? | | | | | |

*0 = Never 1 = Almost never 2 = Sometimes 3 = Fairly often 4 = Very often*

#### LIFESTYLES AND BEHAVIORS

**57. Do you smoke cigarettes (including electronic cigarettes)?***
|__| I don't smoke, or have smoked less than 100 cigarettes in my lifetime, and I don't currently smoke
|__| I am a former-smoker (I have smoked at least 100 cigarettes in my lifetime, but I do not smoke anymore)
|__| Yes, smoking less than 10 cigarettes per day
|__| Yes, smoking between 10 and 20 cigarettes per day
|__| Yes, smoking more than 20 cigarettes a day (more than one pack a day)

*To be completed only if former smoker*

**58. At what age did you quit smoking?*** __________ years
**59. At what age did you start smoking?*** ___________ years
**60. How many cigarettes a day did you smoke before quitting?***
|__| I smoked less than 10 cigarettes a day
|__| I smoked between 10 and 20 cigarettes a day
|__| I smoked more than 20 cigarettes a day (more than one pack a day)

**61. Why did you quit smoking?***
|__| Health problems
|__| Doctor's advice
|__| Personal choice
|__| Illness of relatives or friends

*To be completed only if current smoker*

**62. At what age did you start smoking?*** __________ years

**63. Have you ever smoked or are you currently smoking electronic cigarettes?***
|__| No
|__| Yes, exclusively electronic cigarettes
|__| Yes, not exclusively electronic cigarettes (including tobacco cigarettes)

**64. Does someone usually smoke in your presence in closed environments (at home, in the car, etc.)***
|__| No
|__| Yes, one person
|__| Yes, more than one person

**65. Thinking about the last year, if you have consumed alcohol with meals or between meals, please indicate the frequency***
|__| I don't drink alcohol
|__| Once a month
|__| 2-4 times a month
|__| 2-3 times a week
|__| 4-5 times a week
|__| 6 or more times a week

**66. Since the pandemic started (from February 2020), please indicate if (and how) the following habits or conditions have changed***

| | Not present or not applicable | Decreased | Unchanged | Increased |
|---|---|---|---|---|
| Snacking habits between meals | | | | |

| |
|---|
| Skip one of the main meals |
| Daily consumption of fruit and vegetables |
| Eating a balanced diet (including healthy ingredients such as whole grains, legumes, eggs, nuts, fruits and vegetables) |
| Consumption of sugary foods, drinks, ice cream, sweets, candies |
| Consumption of fried food/fast food |
| Daily/weekly alcohol consumption (during and/or away from meals) |
| Physical activity involving an increase in heart rate and breathing (brisk walking, cycling, swimming, running) |
| Time spent sitting and in front of the screen (TV, computer, mobile phone) |
| Hours of sleep |
| Access to the National Health Service (SSN) for visits and examinations for minor health problems (preventive visits or checks) |
| Access to the National Health Service (SSN) for visits and examinations for major health problems (oncologic, major surgeries, withdrawing of chronic life-sustaining treatments, heart failure, BPCO) |
| Contact with the general practitioner |

#### HOUSING CONDITIONS

**67. What type of heating do you mainly use in your home?***

|__| None
|__| Radiator
|__| Panels (floor, walls)
|__| Fan-coil/split/heat pump/fan coil units
|__| Gas stove
|__| Electric stove
|__| Kerosene stove
|__| Pellet stove
|__| Fireplace/stove or wood boiler

**68. How many rooms are there in your home (excluding the bathroom and auxiliary spaces)?***

|__| One |__| Two |__| Three |__| More than three

**69. Besides you, how many people live in your household?***

|__| None |__| One |__| Two |__| More than two

**70. Are there minors (<18 years old) among your cohabitants?***

|__| No
|__| Yes, one
|__| Yes, two
|__| Yes, three or more

**71. The house you currently live in is ***

|__| Owned |__| Rented |__| Owned by relatives |__| Other (social housing, cooperative housing)

**72. How many cars does your household own?***

0 |__| 1|__| 2 |__| 3 or more |__|

#### SOURCES OF INFORMATION ON COVID-19

**73. Do you feel sufficiently informed about the risks of COVID-19?**
|__| No |__| Yes

**74. What sources of information have you consulted / listened about the risks of COVID-19 and how reliable do you consider them?**

| **Information source** | **No** | **Yes, and I don't trust it** | **Yes, and I partially trust it** | **Yes, and I trust it** |
|---|---|---|---|---|
| Television (national and/or local) | | | | |
| Newspapers and magazines | | | | |
| Radio (national and/or local) | | | | |
| Government, Ministries, Departments, Civil Protection, Law Enforcement, European Union | | | | |
| Newspaper, TV or radio websites | | | | |
| Scientists, researchers, academics | | | | |
| Scientific publications online | | | | |
| Associations or trade unions (environmentalists, volunteers) | | | | |
| Religious institutions | | | | |
| Search engines (Google) | | | | |
| Social media channels (Facebook, Twitter, Instagram, WhatsApp) | | | | |

**75. Below are listed some problems you may encounter in everyday life. What do you think is risky? Is it low, medium or high risk?**

| **Problems** | **Low** | **Medium** | **High** | **I do not know** |
|---|---|---|---|---|
| Natural disasters (earthquakes, floods, etc.) | | | | |
| Unemployment | | | | |
| Pollution of the environment | | | | |
| Epidemics | | | | |
| Poverty | | | | |
| Terrorism | | | | |
| Climate changes | | | | |
| Crime | | | | |
| Contaminated food | | | | |

**Any additional information, reports and comments**

______________________________________________________________________
______________________________________________________________________
____________________________________________________

#### QUESTIONNAIRE EPICOVID19 – PHASE 2

**EXPRESSION OF INTEREST**
Do you agree to be contacted to participate in an Italian study promoted by the National Research Council, which provides for free swabs and the collection of a biological sample (saliva/a few drops of blood)? Among those who join, a random sample will be selected to participate in the national study.
|__| No
|__| Yes

Do you accept to be contacted for future phases of the EPICOVID19 study?
|__| No
|__| Yes

If you agree, please let us know if you want to be contacted at another e-mail address other than the one we already used to contact you back

@______________________________________________________

and / or a telephone number
________________________________
