## Supplementary Tables S1-S5 for "Epidemiology of SARS-CoV-2 infection in Italy using real-world data: methodology and cohort description of the second phase of web-based EPICOVID19 study"

**Table S1.** Contents of the web-based questionnaires.

| **Questionnaires’ sections** | **EPICOVID19 phase I** | **EPICOVID19 phase II** |
| --- | --- | --- |
| ***Socio-demographic characteristics*** |  |  |
| Date of birth | ✓ |  |
| Sex | ✓ | ✓ |
| Country and province of residency | ✓ | ✓ |
| Ethnicity | ✓ |  |
| Educational attainment | ✓ |  |
| Occupational level | ✓ | ✓ |
| Employment status | ✓ | ✓ |
| ***Clinical features and COVID-19 related variables*** |  |  |
| Weight |  | ✓ |
| Height |  | ✓ |
| Blood type |  | ✓ |
| COVID-19-like symptoms | ✓ | ✓ |
| Presence of chronic diseases or conditions | ✓ | ✓ |
| Use of drugs or supplement | ✓ | ✓ |
| Vaccines | ✓ | ✓ |
| COVID-19 vaccine |  | ✓ |
| Use of birth pills and/or hormone replacement therapy | ✓ | ✓ |
| Currently and past pregnancy | ✓ | ✓ |
| Anti-androgens hormone therapy |  | ✓ |
| Contacts with COVID-19 cases | ✓ | ✓ |
| Fiduciary isolation (or quarantine) |  | ✓ |
| COVID-19 nasopharingeal swab testing | ✓ | ✓ |
| Date of COVID-19 nasopharingeal swab testing |  | ✓ |
| Type of COVID-19 nasopharingeal swab testing |  | ✓ |
| Reasons for COVID-19 nasopharingeal swab testing |  | ✓ |
| Time from symptoms |  | ✓ |
| COVID-19 serological testing |  | ✓ |
| Date of COVID-19 serological testing |  | ✓ |
| Type of COVID-19 serological testing |  | ✓ |
| Reasons for COVID-19 serological testing |  | ✓ |
| Contact with emergency number | ✓ | ✓ |
| Hospitalization for COVID-19 | ✓ | ✓ |
| ***Personal characteristics and behaviours*** |  |  |
| Self-reported health status | ✓ | ✓ |
| Fear of SARS-CoV-2^ infection | ✓ | ✓ |
| Sleep disorders |  | ✓ |
| Anxiety and stress perceived |  | ✓ |
| Change of behaviours during the confinement |  | ✓ |
| Population density in the area of residence | ✓ |  |
| Traffic intensity | ✓ |  |
| Number of co-habitants | ✓ | ✓ |
| Number of rooms | ✓ | ✓ |
| Home heating system |  | ✓ |
| Presence of at-risk co-habitants | ✓ |  |
| Presence of co-habitants aged less than 18 years |  | ✓ |
| Daily mean number of contacts | ✓ |  |
| Smoking habit | ✓ | ✓ |
| Alcoholic beverages |  | ✓ |
| Physical activity | ✓ |  |
| Job condition | ✓ |  |
| Going out weekly | ✓ |  |
| Use of public transportation | ✓ |  |
| House property |  | ✓ |
| Cars property |  | ✓ |
| Number of cars |  | ✓ |
| COVID-19 sources of information |  | ✓ |
| Risk perceived |  | ✓ |

^SARS-CoV-2: severe acute respiratory syndrome coronavirus 2.

**Table S2.** Validated scales and questionnaires used in EPICOVID19 survey.

| **Validated scales and questionnaires** | **Measures** |
| --- | --- |
| Educational attainment | http://uis.unesco.org/sites/default/files/documents/international-standard-classification-of-education-isced-2011-en.pdf |
| Occupational status | https://www.eurofound.europa.eu/surveys/ewcs/2005/classification |
| Self-perceived health status | https://ec.europa.eu/eurostat/statistics-explained/index.php?title=Glossary:Self-perceived_health |
| Risk perception | Kim, J.S.; et al. Middle East respiratory syndrome-related knowledge, preventive behaviours and risk perception among nursing students during outbreak. J Clin Nurs. 2016 Sep;25(17-18):2542-9. |
| Clinical features | World Health Organization & International Severe Acute Respiratory and emerging Infection Consortium. (‎2020)‎. Global COVID-19 Clinical Platform Novel Coronavirus (COVID-19)- Rapid Version, “https://www.who.int/docs/default-source/coronaviruse/who-ncov-crf.pdf?sfvrsn=84766e69_2.,” [Online].  Case Record Form Instructions Severe Acute Respiratory Infection Clinical Characterization Data Tool, “ https://media.tghn.org/medialibrary/2019/06/SPRINT-SARI_CRF.V2_Dec_2017__Complete.pdf,” [Online].  M. E. R. S. C. (MERS-CoV), “https://www.who.int/csr/disease/coronavirus_infections/MERS_case_investigation_questionnaire.pdf?ua=1.,” [Online]. |
| Self-reported traffic intensity | Cesaroni, G.; Badaloni, C.; Porta, D.; Forastiere, F.; et al. Comparison between various indices of exposure to traffic-related air pollution and their impact on respiratory health in adults. Occup Environ Med. 2008 Oct;65(10):683-90. |
| Physical activity | Centers for Disease Control and Prevention, “Strategies to Prevent Obesity and Other Chronic Diseases: The CDC Guide to Strategies to Increase Physical Activity in the Community. Atlanta: U.S. Department of Health and Human Services; 2011. |
| Sleeping disorders | Jenkins, C.D.; Stanton, B.A.; Niemcryk, S.J.; Rose, R.M. A scale for the estimation of sleep problems in clinical research. J Clin Epidemiol 1988;41(4):313-21 |
| Smoking habit | National Health Interview Survey – Adult tobacco use information. Centers for Disease Prevention and Control. 2017 Aug  29 https://www.cdc.gov/nchs/nhis/tobacco/tobacco_glossary.htm |
| Perceived stress | Cohen, S.; Kamarck, T.; and Mermelstein, R. A global measure of perceived stress. Journal of Health and Social Behavior. 1983, 24, 386-396 (adapted) |
| Alcohol consumption | Frank, D.; De Benedetti, A.F.; Volk, R.J.; Williams, E.C.; Kivlahan, D.R.; Bradley, K.A. [Effectiveness of the AUDIT-C as a screening test for alcohol misuse in three race/ethnic groups](https://doi.org/10.1007/s11606-008-0594-0). J Gen Intern Med. 2008, 23(6):781-787 (adapted) |
| Deprivation index | Townsend, P.; Phillimore, P.; Beattie, A. (1988) Health and Deprivation: Inequality and the North Croom Helm: London |

**Table S3.** Characteristics of individuals who participated in phase II survey (N=41,473) and of those who did not (N=157,349).

|  |  | **Phase II survey** | |  |
| --- | --- | --- | --- | --- |
|  |  | **Participated**  **(N=41,473)** | **Did not participate**  **(N=157,349)** | ***p-value*** |
| Sex | Females | 25,146 (60.6) | 93,524 (59.4) |  |
|  | Males | 16,327 (39.4) | 63,825 (40.6) | <.001 |
| Class of age | 18-29 | 3,255 (7.8) | 21,564 (13.7) |  |
|  | 30-39 | 7,336 (17.7) | 30,427 (19.3) |  |
|  | 40-49 | 9,274 (22.4) | 33,468 (21.3) |  |
|  | 50-59 | 10,616 (25.6) | 35,415 (22.5) |  |
|  | 60-69 | 8,217 (19:8) | 25,416 (16.2) |  |
|  | 70-79 | 2,523 (6.1) | 9,083 (5.8) |  |
|  | 80+ | 252 (0.6) | 1,976 (1.3) | <.001 |
| Italian area | Northern | 29,072 (70.1) | 104,605 (66.5) |  |
|  | Central | 8,282 (20.0) | 32,600 (20.7) |  |
|  | Southern and islands | 4,096 (9.9) | 19,754 (12.6) |  |
|  | Abroad or unknown | 23 (0.1) | 390 (0.2) | <.001 |
| Educational level | Low | 1,381 (3.3) | 10,473 (6.7) |  |
|  | Middle | 12,934 (31.2) | 55,866 (35.5) |  |
|  | High | 27,158 (65.5) | 91,010 (57.8) | <.001 |
| Education | None | 19 (0.0) | 253 (0.2) |  |
|  | Elementary school | 36 (0.1) | 987 (0.6) |  |
|  | Secondary school license | 1,326 (3.2) | 9,233 (5.9) |  |
|  | Secondary school diploma | 12,934 (31.2) | 55,866 (35.5) |  |
|  | University degree | 18,263 (44.0) | 63,666 (40.5) |  |
|  | Postgraduate degree | 8,895 (21.4) | 27,344 (17.4) | <.001 |
| Employment status | Employed | 29,299 (70.6) | 107,557 (68.4) |  |
|  | Student | 1,703 (4.1) | 11,733 (7.5) |  |
|  | Unemployed | 1,671 (4.0) | 7,513 (4.8) |  |
|  | Retired | 6,611 (15.9) | 21,697 (13.8) |  |
|  | Other | 2,189 (5.3) | 8,849 (5.6) | <.001 |
| Healthcare workers | No | 38,179 (92.1) | 145,907 (92.7) |  |
|  | Yes | 3,294 (7.9) | 11,442 (7.3) | <.001 |
| N° of morbidities | None | 25,466 (61.4) | 101,791 (64.7) |  |
|  | One | 11,628 (28.0) | 40,736 (25.9) |  |
|  | Two | 3,356 (8.1) | 11,181 (7.1) |  |
|  | Three or more | 1,023 (2.5) | 3,641 (2.3) | <.001 |
| Number of rooms in the house | One | 414 (1.0) | 1,752 (1.1) |  |
|  | Two | 3,998 (9.6) | 15,583 (9.9) |  |
|  | Three | 10,596 (25.5) | 40,403 (25.7) |  |
|  | More than three | 26,465 (63.8) | 99,611 (63.3) | .049 |
| Number of cohabitant people | No one | 5,713 (13.8) | 19,383 (12.3) |  |
|  | One | 13,819 (33.3) | 49,205 (31.3) |  |
|  | Two | 9,611 (23.2) | 37,699 (24.0) |  |
|  | More than two | 12,330 (29.7) | 51,062 (32.5) | <.001 |

**Table S4.** Standardized response rates per 100,000 inhabitants by Italian region over the phase II study period.

| **Region** | **Frequency** | **Percentage** | **Italian population*** | **Response rate** |
| --- | --- | --- | --- | --- |
| Abruzzo | 330 | 0.8 | 1,311,580 | 25.2 |
| Basilicata | 110 | 0.3 | 562,869 | 19.5 |
| Calabria | 199 | 0.5 | 1,947,131 | 10.2 |
| Campania | 886 | 2.1 | 5,801,692 | 15.3 |
| Emilia-Romagna | 4,485 | 10.8 | 4,459,477 | 100.6 |
| Friuli-Venezia Giulia | 812 | 2.0 | 1,215,220 | 66.8 |
| Lazio | 4,072 | 9.8 | 5,879,082 | 69.3 |
| Liguria | 1494 | 3.6 | 1,550,640 | 96.3 |
| Lombardia | 13,832 | 33.4 | 10,057,370 | 137.5 |
| Marche | 710 | 1.7 | 1,525,271 | 46.5 |
| Molise | 63 | 0.2 | 305,617 | 20.6 |
| Piemonte | 4,644 | 11.2 | 4,356,406 | 106.6 |
| Puglia | 918 | 2.2 | 4,029,053 | 22.8 |
| Sardegna | 868 | 2.1 | 1,639,591 | 52.9 |
| Sicilia | 722 | 1.7 | 4,999,891 | 14.4 |
| Toscana | 3,188 | 7.7 | 3,729,641 | 85.5 |
| Trentino | 598 | 1.4 | 1,072,276 | 55.8 |
| Umbria | 312 | 0.8 | 882,015 | 35.4 |
| Valle d'Aosta | 76 | 0.2 | 125,666 | 60.5 |
| Veneto | 3,131 | 7.5 | 4,905,854 | 63.8 |

*Resident in Italy on January 1^st^, 2020 (ISTAT)

**Table S5.** Frequency of self-reported symptoms by sex and by positivity to NPS and/or ST between March 2020 and February 2021 (N=41,473).

| **Self-reported symptoms** | **Total**  **(N=41,473)** | **Total positive cases**  **(N=4,411)** | **Females**  **(N=25,146)** | **Females positive**  **cases (N=2,716)** | **Males (N=16,327)** | **Males positive cases (N=1,695)** |
| --- | --- | --- | --- | --- | --- | --- |
| Fever | 6,290 (15.2) | 2,560 (58.0) | 3,755 (14.9) | 1,504 (55.4) | 2,535 (15.5) | 1,056 (62.3) |
| Cough | 6,159 (14.9) | 1,908 (43.3) | 3,672 (14.6) | 1,142 (42.1) | 2,487 (14.9) | 766 (45.2) |
| Headache | 11,554 (27.9) | 2,324 (52.7) | 8,027 (31.9) | 1,578 (58.1) | 3,527 (21.6) | 746 (44.0) |
| Myalgia | 9,064 (21.9) | 2,849 (64.6) | 6,066 (24.1) | 1,837 (67.6) | 2,998 (18.4) | 1,012 (59.7) |
| Shortness of breath | 3,272 (7.9) | 1,364 (30.9) | 2,134 (8.5) | 893 (32.9) | 1,138 (7.0) | 471 (27.8) |
| Chest pain | 2,054 (5.0) | 731 (16.6) | 1,346 (5.4) | 495 (18.2) | 708 (4.3) | 236 (13.9) |
| Gastrointestinal | 7,330 (17.7) | 1,553 (35.2) | 4,840 (19.2) | 1,047 (38.6) | 2,490 (15.3) | 506 (29.9) |
| Sore throat/rhinorrhoea | 10,157 (24.5) | 1,927 (43.7) | 6,493 (25.8) | 1,256 (46.2) | 3,664 (22.4) | 671 (39.6) |
| Dysgeusia | 3,299 (8.0) | 2,129 (48.3) | 2,205 (8.8) | 1,418 (52.2) | 1,094 (6.7) | 711 (42.0) |
| Anosmia | 3,391 (8.2) | 2,275 (51.6) | 2,280 (9.1) | 1,518 (55.9) | 1,111 (6.8) | 757 (44.7) |
| Loss of appetite | 1,778 (4.3) | 932 (21.1) | 1,229 (4.9) | 639 (23.5) | 549 (3.4) | 293 (17.3) |
| Cardiological | 2,732 (6.6) | 679 (15.4) | 1,951 (7.8) | 499 (18.4) | 781 (4.8) | 180 (10.6) |
| Dermatological | 2,304 (25.6) | 456 (10.3) | 1,534 (6.1) | 316 (11.6) | 770 (4.7) | 140 (8.3) |
| Neurological | 1,747 (4.2) | 648 (14.7) | 1,285 (5.1) | 490 (18.0) | 462 (2.8) | 158 (9.3) |

Number are counts and row percentages
